# Clinical validity and utility of circulating tumor DNA (ctDNA) testing in advanced non-small cell lung cancer (aNSCLC): a systematic literature review and meta-analysis

**DOI:** 10.1101/2023.10.27.23297657

**Authors:** Cheng Chen, Michael P. Douglas, Meera V. Ragavan, Kathryn A. Phillips, Jeroen P. Jansen

## Abstract

**Purpose:** Circulating tumor DNA (ctDNA) testing has become a promising tool to guide first-line (1L) targeted treatment for advanced non-small cell lung cancer (aNSCLC). This study aims to estimate the clinical validity (CV) and clinical utility (CU) of ctDNA-based next-generation sequencing (NGS) for oncogenic driver mutations to inform 1L treatment decisions in aNSCLC through a systematic literature review and meta-analysis.

**Methods:** A systematic literature search was conducted in PubMed/MEDLINE and Embase to identify randomized control trials or observational studies reporting CV/CU on ctDNA testing in patients with aNSCLC. Meta-analyses were performed using bivariate random-effects models to estimate pooled sensitivity and specificity. Progression-free/overall survival (PFS/OS) was summarized for CU studies.

**Results:** Eighteen studies were identified: 17 CV only, 2 CU only, and 1 both. Thirteen studies were included for the meta-analysis on multi-gene detection. The overall sensitivity and specificity for ctDNA detection of any mutation were 0.69 (95% CI, 0.63–0.74) and 0.99 (95% CI, 0.97–1.00) respectively. However, sensitivity varied greatly by driver gene, ranging from 0.29 (95% CI, 0.13–0.53) for *ROS*1 to 0.77 (95% CI, 0.63–0.86) for *KRAS*. Two studies compared PFS with ctDNA versus tissue-based testing followed by 1L targeted therapy found no significant differences. One study reported OS curves on ctDNA-matched and tissue-matched therapies but no hazard ratios were provided.

**Conclusion:** ctDNA testing demonstrated an overall acceptable diagnostic accuracy in aNSCLC patients, however, sensitivity varied greatly by driver mutation. Further research is needed, especially for uncommon driver mutations, to better understand the CU of ctDNA testing in guiding targeted treatments for aNSCLC.

## Introduction

Non-small cell lung cancer (NSCLC) accounts for an estimated 85% of lung cancer cases, with adenocarcinoma, squamous cell carcinoma, and large cell carcinoma being the most common histological subtypes.^1^ The estimated five-year survival is only about 6% in patients with advanced-stage disease.^2,3^ In the past decade, more than 20 targeted therapies have been approved for the treatment of advanced NSCLC (aNSCLC) in patients who harbor *EGFR*, *BRAF*, *MET*, *RET*, *NTRK*, *KRAS*, *ALK*, or *ROS1* alterations.^4^ Targeted treatment according to the presence of oncogenic driver mutations has been associated with improved survival outcomes^4–6^, and current National Comprehensive Cancer Network (NCCN) guidelines specifically recommend that all patients with aNSCLC should get broad genomic profiling with next-generation sequencing (NGS), given its more optimal use of sample availability, reduced procedure time, and favorable testing costs compared to single gene tests.^7–9^

In recent years, there has been an increase in the use of circulating tumor DNA (ctDNA) from blood samples as an alternative to tissue biopsy (TB) for identifying oncogenic driver mutations to inform first-line (1L) aNSCLC therapy.^10^ Advantages of ctDNA testing include the avoidance of an invasive procedure and possible complications, the ability to identify driver mutations for patients with limited tissue availability for comprehensive NGS, and shorter turnaround time allowing for faster initiation of 1L therapy.^11^ Several ctDNA-based NGS tests have been developed and approved for NSCLC, including Guardant360^®^ CDx and FoundationOne^®^Liquid CDx.^12,13^

However, the diagnostic accuracy of ctDNA testing remains unclear due to variations in technologies and use scenarios.^14,15^ With the broadened adoption of ctDNA-based NGS tests in NSCLC, a better understanding of their clinical validity (CV) and clinical utility (CU) is needed. “Clinical validity” usually describes the diagnostic accuracy of genetic tests and is generally measured by sensitivity, specificity, positive predictive value (PPV), and negative predictive value (NPV).^16,17^ Without proper validation of test performance, accurate identification of mutations may be compromised, leading to delays in suitable clinical decisions. Previous systematic literature reviews have examined the CV of ctDNA testing in aNSCLC patients but findings have been inconsistent due to varied review focus and inclusion criteria.^18–26^ Most of the previous systematic literature reviews and/or meta-analyses^18,19,22,23,25,26^ did not specify sequencing technologies, which could potentially explain the observed heterogeneity in findings and limit their clinical implications in relation to the currently more recommended NGS tests.

“Clinical utility” refers to the risks or benefits from test use and in our study is measured as progression-free or overall survival (PFS/OS), considering the therapeutic effectiveness outcomes are the most relevant and commonly reported measure in CU studies and that the focus of our review is 1L targeted treatment informed by ctDNA testing.^27–29^ CU evidence directly answers if a test is useful in improving patient health outcomes – it is an essential element in value evaluation and affects the acceptance of a test from all parties (patients, healthcare providers and payers). To our knowledge, no study has systematically reviewed the CU of ctDNA testing for a comprehensive set of biomarkers to inform 1L treatment decisions in aNSCLC.

Given the rapidly evolving field of ctDNA testing technologies and the limitations of previous review studies, an updated synthesis of CV and CU evidence is warranted. The objective of the current study was to estimate the CV and CU of ctDNA-based NGS for oncogenic driver mutations to inform 1L treatment decisions in aNSCLC by means of a systematic literature review and meta-analysis of currently available evidence.

## Materials and Methods

### Systematic literature review

#### Eligibility criteria

The study inclusion criteria were defined in terms of the population, interventions, comparisons, outcomes, and study design (PICOS) to guide the identification and selection of relevant studies: *Population* - Adult patients with aNSCLC (at least 80% with stage III or IV thereby making the assumption that reported results based on the total study population are still applicable to the stage III or IV NSCLC target population of interest), with a subset of treatment-naïve patients for CV studies and all being treatment-naïve for CU studies; *Interventions* – ctDNA-based NGS especially for the detection of clinically relevant driver mutations (i.e., *EGFR*, *BRAF*, *MET*, *RET*, *NTRK*, *KRAS*, *ALK*, *ROS1*), and when outcome of interest is CU at least 80% of population receiving matched targeted treatment by driver mutation according to current NCCN guidelines^7^; *Comparators for CU studies* – ctDNA testing vs. TB; *Outcomes in CV studies* – sensitivity, specificity, PPV, NPV or any other measures that allow calculation of true positive (TP), false positive (FP), false negative (FN), and true negative (TN) rates; *Outcomes in CU studies* – PFS or OS. *Study design* – cohort studies or randomized clinical trials. Studies were excluded if the study population excluded patients with any of the eight clinically relevant driver mutations; published in a non-English language; published before 2012 (one year before the first FDA approval of liquid biopsy test); and were review articles or conference proceedings.

#### Study identification

Relevant studies published between January 2012 and July 2023 were identified by searching MEDLINE and Embase databases with predefined search strategies (Supplementary File, Table S2) through the Embase platform. Furthermore, the official websites of three commonly used ctDNA tests (Guardant360^®^ CDx, InvisionFirst^®^-Lung, and FoundationOne^®^ Liquid CDx) as well as reference lists of included studies and previous systematic literature reviews^18–26^ were searched for additional potentially eligible studies.

#### Study selection

Two reviewers (CC and MD) screened the identified abstracts using an open-source, active learning software/platform, ASReview, following recommended approaches for automated screening.^30–34^ Studies identified as eligible during abstract screening were subsequently screened at a full-text stage by the same two reviewers according to the eligibility criteria to determine the final set of included studies. Following reconciliation between the two investigators, a third reviewer (JJ) was included to reach a consensus for any remaining discrepancies. The process of study identification and selection was summarized with a Preferred Reporting Items for Systematic Reviews and Meta-Analyses (PRISMA) flow diagram.

#### Data extraction

Two reviewers (CC and MD) extracted data on study characteristics, test and intervention characteristics, patient characteristics, and outcomes for the final list of included studies. Data was stored and managed in a Microsoft Excel workbook. *Trial characteristics* - author name, publication year, country, sample size, oncogenic driver mutations of interest, study duration, and patient in/exclusion criteria; *patient characteristics* - disease stage, smoking status, race/ethnicity, gender, and age; *test and intervention characteristics -* ctDNA and NGS technologies and in addition for CU studies, targeted treatment regimen; *Outcomes in CV studies –* overall and oncogenic driver mutation-specific sensitivity, specificity, TP, FP, FN, TN; *Outcomes in CU studies* - PFS and/or OS.

#### Quality assessment

Two authors (CC and MD) independently assessed the quality of included CV studies based on the revised Quality Assessment of Diagnostic Accuracy Studies-2 (QUADAS-2) criteria.^35^ Discrepancies were resolved by consensus.

#### Analysis

Study-specific, sample (mutation)-level TP, FP, FN, and TN frequency data for oncogenic driver mutations were used to estimate the overall and mutation-specific sensitivity and specificity for each study as well as across studies by meta-analyses. The overall sensitivity and specificity were estimated based on studies that simultaneously tested at least four driver mutations. Study-specific sensitivity and specificity estimates along with 95% confidence intervals (95% CIs) were obtained according to the Wilson method using continuity corrected cell counts.^36,37^ The bivariate random-effects model proposed by Reitsma et al. (2005) utilizes the standard frequentist approach and was used in our study to obtain pooled estimates for sensitivity and specificity.^38–40^ Meta-analysis results were also presented with summary receiver operating characteristics (SROC) curves and 95% confidence regions for the pooled sensitivity and specificity estimates. In addition, as a sensitivity analysis, a Bayesian bivariate random-effects meta-analysis was performed to avoid normal approximations of the likelihood and to obtain predictive distributions of the sensitivity and specificity to predict results in a new study. Both the frequentist and Bayesian approaches used in our study modeled the sum and differences of true positive and false positive rates as random effects. The Bayesian receiver operating characteristics (BSROC) curves and Bayesian area under the BSROC curve (BAUC) were reported with 95% CI for the ctDNA detection of any and each driver mutation. An AUC of 0.7 or higher is generally considered good accuracy and 0.6-0.7 is considered sufficient; an AUC below 0.5 indicates the test is not useful.^41^ There is no standard for good sensitivity/specificity of DNA testing. A ctDNA test might be considered acceptable for coverage if its performance is similar to the FDA-approved Guardant360^®^.^42^ All analyses were conducted using RStudio, version 4.1.2 (©2009-2022 RStudio, PBC) using packages “mada” and “bamdit”.^38,43^ Progression-free/overall survival (PFS/OS) was summarized for CU studies.

## Results

### Study selection

Our initial search generated 1,749 potentially relevant publications. After screening titles and abstracts, 58 publications were selected for full-text review. A total of 20 publications corresponding to 20 studies were selected for inclusion (Figure 1). Sixteen studies^44–59^ reported CV only, one study^60^ reported CU only, and one study^61^ provided information on both CV and CU.

**Figure 1.**
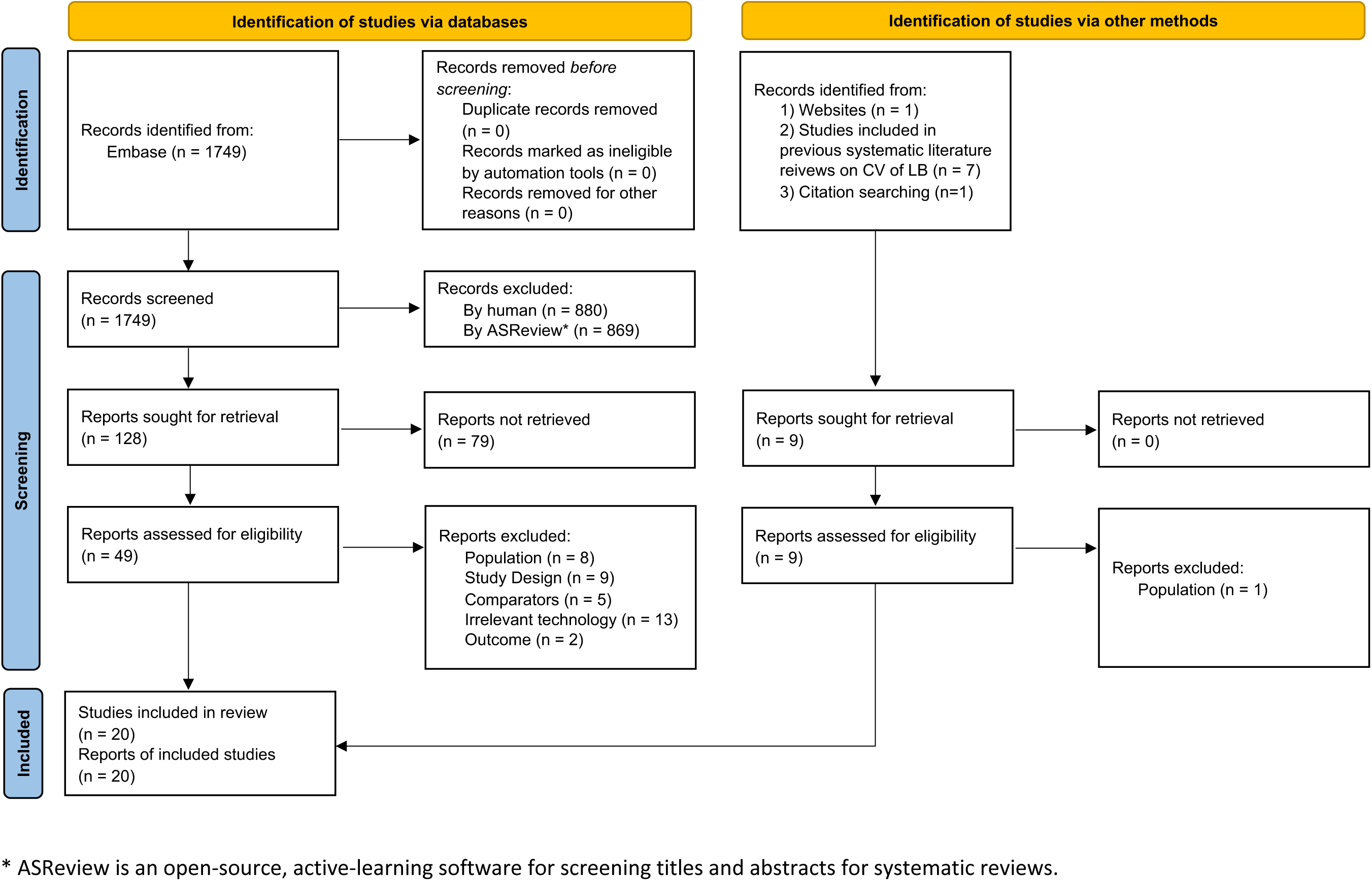
The PRISMA flowchart of included studies

### Clinical validity of ctDNA testing to identify oncogenic driver mutations

#### Characteristics of included studies

The majority of the included CV studies^45–47,49–51,53–59,61,62^ used a prospective cohort design (83.3%, 15/18), and three studies^44,48,52^ used retrospective medical record data. All studies evaluated ctDNA-based NGS technologies, with eleven studies^44–46,48–52,58,61,62^ evaluating branded ctDNA tests and seven studies^47,53–57,59^ describing the ctDNA technologies without a specific brand name (Table 1). Guardant360^®^ was the most used ctDNA technology (33.3%, 6/18). TB was the reference standard in all included studies.

**Table 1.** Characteristics of included CV studies (n=18)

| Author_Year | Country | No. Pts* | LB Technology | TB Technology | Time between testing (days) | Test genes/driver mutations (included in analysis) |
| --- | --- | --- | --- | --- | --- | --- |
| Bustamante Alvarez_2021 | US | 143 | Guardant360® | Fluorescence in situ hybridization (FISH) | ≤84 | *EGFR*, *ALK* fusions, *BRAF*, *MET* exon 14 skipping mutation, *ROS1* mutations and *RET* fusions; *KRAS* |
| Fernandes_2021 | Portugal | 115 | Oncomine™ Lung cfDNA | Unspecified | -- | Overall Oncomine panel (*EGFR*, *KRAS*, *ALK*, *BRAF*, *MET*, *ROS1*, *ERBB2*, *MAP2K1*, *NRAS*, *PIK3CA*, and *TP53*) |
| Leighl_2019 | US | 282 | Guardant360® | Unspecified | -- | *EGFR*, *ALK* fusion, *BRAF* V600E, *MET* amplification, *MET* exon 14 skipping, *ROS1* fusion, *RET* fusion, *ERBB2* mutation |
| Li_2019 | US | 127 | Unspecified | MSK-IMPACT | 207 (median) | *EGFR*, *KRAS*, *ALK*, *BRAF*, *MET*, *ROS1*, *RET*, *ERBB2* |
| Lin_2021 | US | 100 | Guardant360® | NYU Oncomine™ Focus | 12 (median) | Overall Guardant 360 panel (patient level) |
| Müller_2017 | Germany | 98 | NEOliquid® | NEOPlus, PCR and FISH | -- | *EGFR*, *KRAS*, *ALK*, *BRAF*, *ROS1*, *TP53* |
| Palmero_2021 | Spain | 186 | Guardant360® | Unspecified | -- | *EGFR*, *ALK* |
| Park_2021 | Korea | 421 | Guardant360® | Oncomine™ Focus | ≤1 | *EGFR* Del 9, *EGFR* L858R, *KRAS*, *ALK*, *BRAF* V600E, *MET* exon 14, *MET* amplification, *ROS1*, *RET*, *ERBB2* exon 20 |
| Pritchett_2019 | US | 264 | InVisionFirst® | Custom-designed SureSelect XT assay and FusionPlex Tumor Panel | ≤84 | *EGFR* (exons 18-21), *KRAS*, *ALK/ROS1*, *BRAF* V600E, *MET* exon 14, *ERBB2* exon 20, *STK11* |
| Schwaederlé_2017 | US | 88 | Guardant360® | FoundationOne | -- | Overall Guardant 360 panel (patient level) |
| Yao_2017 | China | 39 | Unspecified | Unspecified | ≤7 | *EGFR*, *KRAS*, *ALK*, *RET*, *PIK3CA* |
| Xu_2016 | China | 42 | Unspecified | Unspecified | -- | 50 gene panel (*EGFR*, *KRAS*, *BRAF*, *ERBB2*, *PIK3CA*, *TP53* identified) |
| Liu_2018 | China | 72 | Unspecified | Unspecified | ≤7 | *EGFR* L858R, *EGFR* exon 19 del, *KRAS* G12X, *ALK* rearrangement |
| Couraud_2014 | France | 106 | Unspecified | Unspecified | -- | *EGFR* (exons 18-21), *KRAS* (exons 2 & 3), *BRAF* (exons 11 & 15), *ERBB2* (exons 19 & 20), *PI3KCA* (exons 9 & 20) |
| Chen_2019 | China | 50 | Unspecified | Unspecified | -- | >200 gene variants; specific CV on *EGFR* |
| Remon_2019 | France | 194 | InVisionFirst® | Unspecified | 34 (median) | *EGFR* (exons 18-21), *KRAS*, *BRAF* V600E, *MET* exon 14, *ERBB2* exon 20, *STK11* |
| Guo_2016 | China | 41 | SV-CA50-ctDNA panel | Ion Proton System, Ion PI sequencing | ≤13 | *EGFR*, *KRAS*, *BRAF*, *ERBB2*, *PIK3CA*, *TP53* |
| Sabari_2019 | US and Australia | 210 | ResBio ctDx-Lung | MSK-IMPACT | 21 (median) | *EGFR*, *KRAS*, *ALK*, *BRAF*, *MET*, *ROS1*, *RET* |

\* Baseline overall patient sample size

Seven studies^45,46,50,51,55,56,61^ included only untreated patients, five studies^44,47,48,53,58^ included 24-92% of untreated patients, and in six studies^49,52,54,57,59,62^ the proportion of untreated patients was unclear. Among studies reporting patient racial/ethnic information^46,51,52,61^, most patients were Caucasian (Table S3). The type of driver mutations examined differed across studies. No study reported CV on all eight clinically relevant driver mutations. Most of the studies evaluated ctDNA detection of EGFR (77.8%, 14/18) and *KRAS* (61.1%, 11/18). Thirteen studies reported CV information on six or more mutations (Table S4).

Four studies were found to have no risk of bias or applicability concerns and nine studies had two or fewer items of concern according to the QUADAS-2 instrument (Table S16). Eight studies were found to have unclear-risk items, indicating potential issues with the reporting of CV evidence.

#### Overall sensitivity and specificity for detection of any guideline-recommended oncogenic driver mutation

Overall sensitivity for the detection of any driver mutation varied between 0.52 and 0.81 across 13 studies that simultaneously detected four or more driver mutations (Figure 2 and Supplementary File Table S4). The overall specificity varied between 0.88 and 1, with 11 studies reported a specificity value of 0.90 or higher. There was a small degree of between-study heterogeneity regarding sensitivity and specificity (*I*^2^=20%).^63^ The pooled sensitivity obtained with meta-analysis using the frequentist approach was 0.69 (95% CI, 0.63–0.74) and the pooled specificity was 0.99 (95% CI, 0.97–1.00). Bayesian meta-analysis generated similar pooled estimates: sensitivity=0.70 (95% CI, 0.60-0.79); specificity=0.99 (95% CI, 0.97-1.00). The SROC curves with both approaches and the 95% confidence regions for the pooled sensitivity and specificity estimates are in Figure 3. The BAUC was 0.71 (95% CI, 0.68-0.73). In general, overall sensitivity was higher in studies that used branded ctDNA tests compared to studies that did not describe tests with a specific brand name, except for the ResBio ctDx_Lung used in the study by Sabari et al. Overall specificity was high regardless of the ctDNA test used. We did not observe a positive relationship between sensitivity/specificity and study quality.

**Figure 2.**
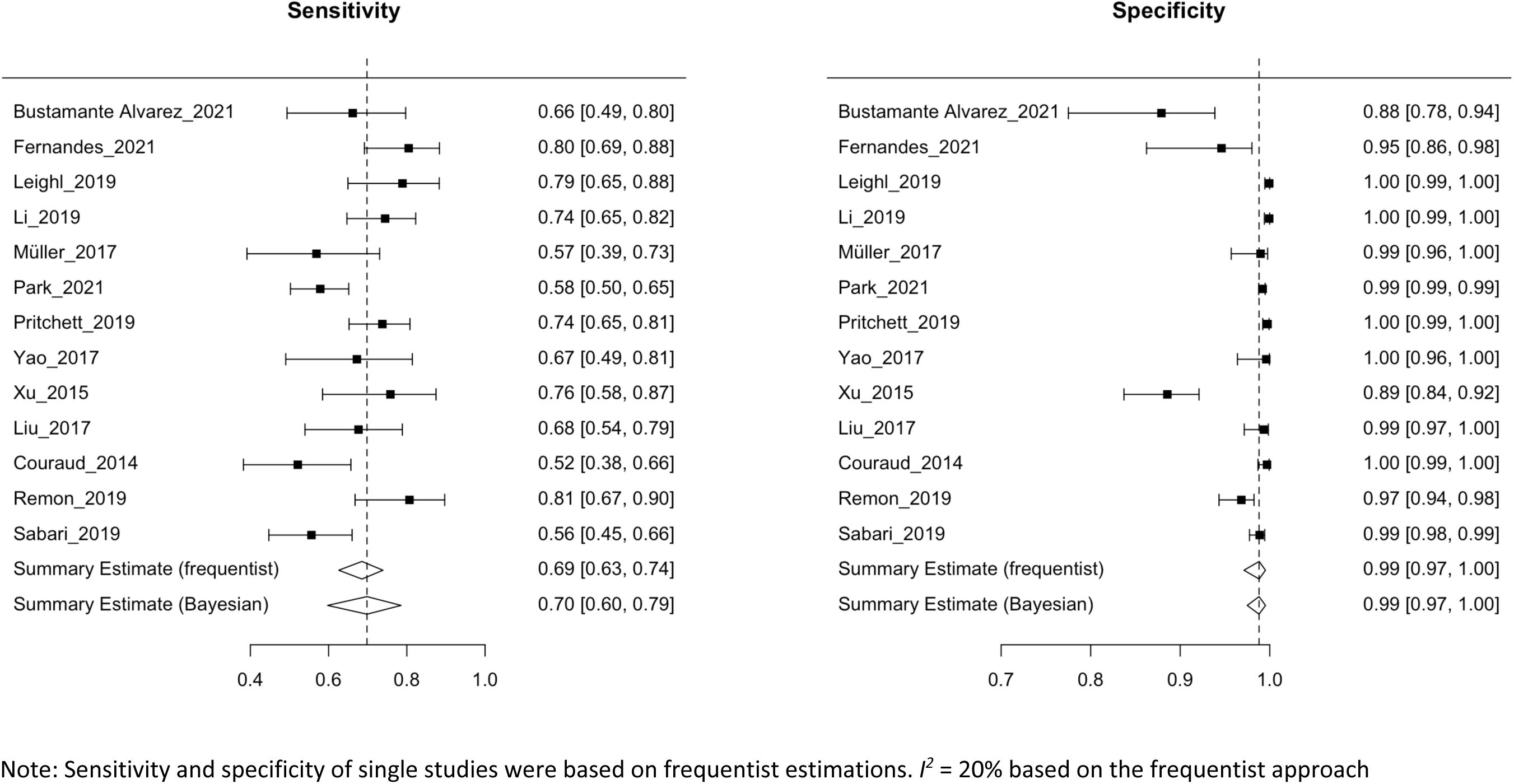
Forest plot of sensitivity and specificity of ctDNA testing for multi-gene detection from bivariate random-effects meta-analyses (n=13)

**Figure 3.**
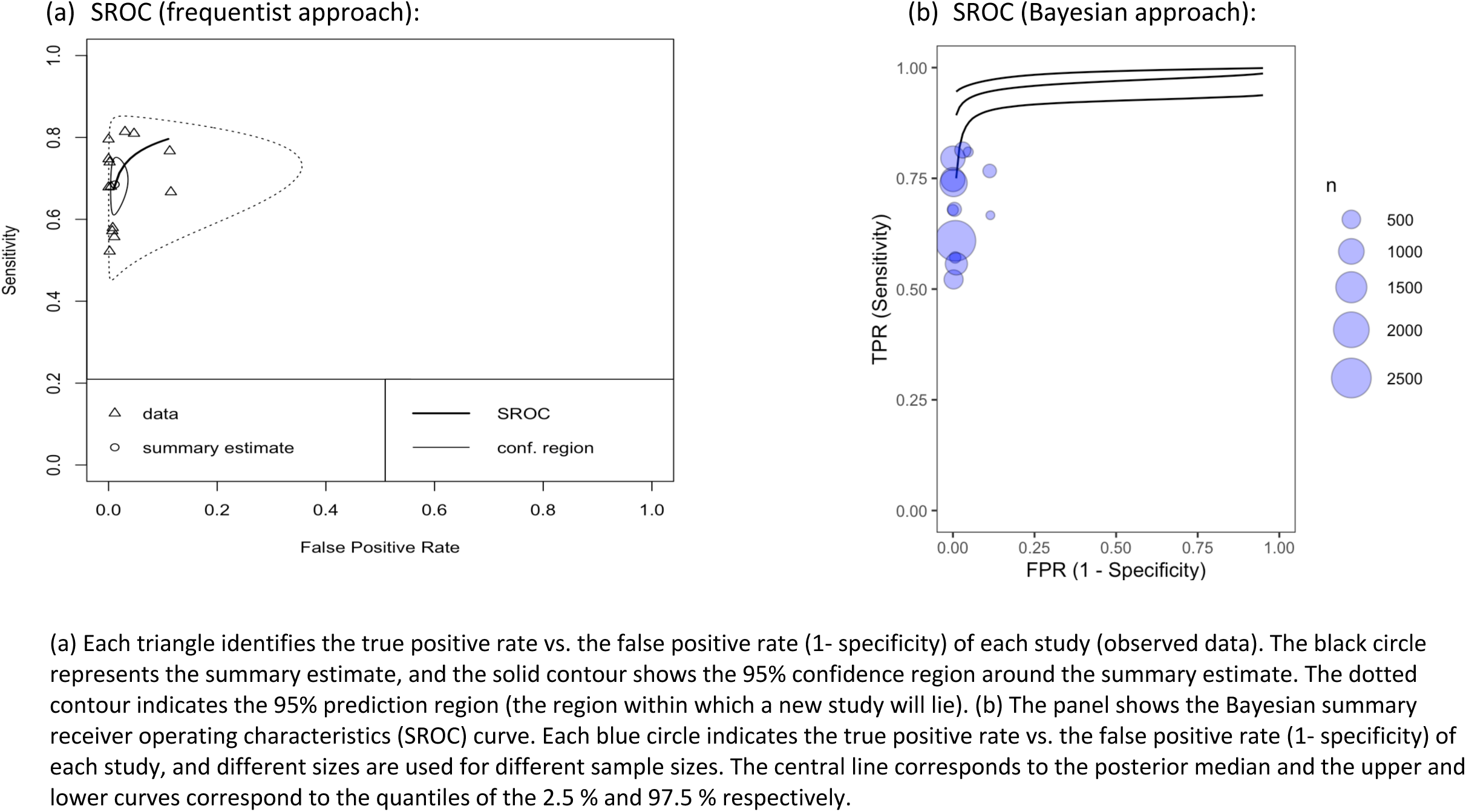
Summary receiver operating characteristics (SROC) plots based on frequentist and Bayesian bivariate random-effects meta-analyses (n=13)

#### Sensitivity and specificity by oncogenic driver mutation

An overview of meta-analysis results of sensitivity and specificity by oncogenic driver mutation is presented in Table 2. Corresponding SROCs and study-specific estimates are provided in Supplementary File, Table S6-S12 and Figure S2-S22. Sensitivity and specificity for detecting *EGFR* with ctDNA were reported in 14 studies.^46,47,49–58,61,62^ Study-specific sensitivity estimates varied between 0.56 and 0.83, and specificity estimates varied between 0.68 and 1 (Figure S2). The pooled sensitivity and specificity as estimated with the meta-analysis were 0.68 and 0.98 respectively, with a BAUC of 0.71 (95% CI, 0.68-0.73). Pooled sensitivity for the other driver mutations was generally less than 0.70 (except 0.77 for *KRAS*), but specificity was close to 1. The pooled sensitivity for each driver mutation detection was generally higher for branded ctDNA tests compared to tests that were not described with a specific brand name; overall pooled specificity was still high regardless of ctDNA tests used. We also conducted an analysis of sensitivity and specificity of ctDNA testing for detecting different mutation classes, including SNVs, indels and fusions. We found that only SNVs are associated with an acceptable BAUC of 0.72 (95% CI, 0.70-0.74), while the other mutation classes demonstrated a BAUC of around 0.52 (Tables S13-S15, Figures S23-S31).

**Table 2.**
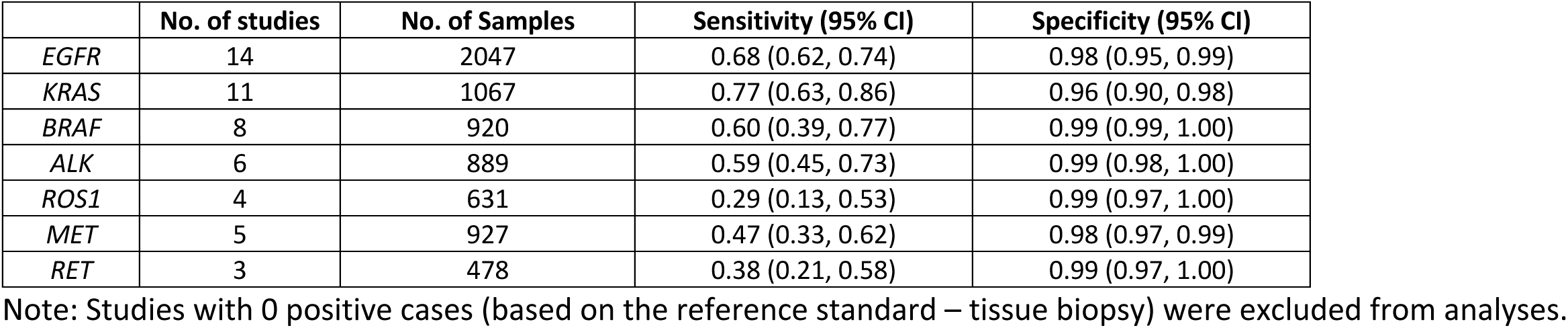
Results from frequentist bivariate random-effects meta-analyses of ctDNA testing for single-gene detection.

### Clinical utility of ctDNA testing to inform 1L treatment in patients with aNSCLC

Three studies evaluated the CU of ctDNA testing to inform 1L targeted therapy.^60,61,64^ Madison *et al.* compared PFS among patients on matched 1L therapies following comprehensive genomic profiling of ctDNA (FoundationOne^®^Liquid or FoundationACT^®^) and TB (FoundationOne^®^CDx or FoundationOne^®^) using data from the Flatiron Health-Foundation Medicine Clinico-Genomic Database.^60^ Median PFS in the ctDNA group (n=33) was 13.8 (95% CI, 8.9-NA) months and 10.6 (95% CI, 8.7-13.6) months in the TB group (n=229). The hazard ratio of ctDNA vs. TB regarding PFS was 0.68 (95% CI, 0.36-1.26) – not statistically significant. OS was only reported according to matched or unmatched 1L treatment but not specifically for TB and ctDNA. Palmero et al. reported PFS for 41 patients treated with 1L targeted therapy informed by ctDNA or TB testing (Guardant360^®^) (median, 8.6; 95% CI, 7.6-11.6 months).^61^ According to the reported Kaplan-Meier (KM) curves, PFS was comparable between these two groups. Jee et al. reported OS for ctDNA-matched (median OS of 39 months) and tissue-matched treatment-naïve patients (29 months), however they did not provide a relative treatment effect estimate independent of whether driver mutations were detected with ctDNA to infer the clinical utility of ctDNA relative to TB testing.^64^

## Discussion

Our meta-analyses showed that ctDNA testing demonstrated acceptable sensitivity and high specificity for detecting any guideline-recommended oncogenic driver mutation. However, the sensitivity of ctDNA testing varied widely by driver mutation. Pooled sensitivity estimates indicated acceptable performance for KRAS, but estimates were less than 70% for the other driver mutations. Specificity of ctDNA testing was high for all mutations. Evidence regarding the CU of ctDNA testing relative to TB was limited. According to a single study, there was no difference in PFS between ctDNA and TB tests followed by 1L targeted therapy.

To our knowledge, this is the first systematic literature review and meta-analysis specifically focused on the diagnostic performance of ctDNA testing for multi-gene detection with NGS in patients with aNSCLC to inform 1L targeted therapy. In contrast to previous systematic literature reviews/meta-analyses on ctDNA detection of EGFR or KRAS^18,22,23,25,26^, we assessed the diagnostic accuracy of ctDNA for detecting multiple driver mutations simultaneously rather than focusing on single mutation thereby providing more relevant information for routine clinical practice where ctDNA-based NGS is rapidly being adopted. Meta-analysis of diagnostic accuracy can be performed based on results reported at the patient level, or the sample (mutation)-level. Previous meta-analyses mostly reported ctDNA performance at the patient level^18,22,23^ or did not specify the data level.^19,20^ We used sample-level data to facilitate diagnostic performance by driver mutation as well as mutation class, which allows for a more detailed assessment of the CV of ctDNA testing and more mutation-specific information.

In our systematic review, we included any study evaluating ctDNA testing for which the study population included at least 80% aNSCLC patients and any proportion of treatment-naïve patients, which did not strictly align with our target patient population of interest – aNSCLC patients initiating 1L treatment. The reason to cast a wider net was to ensure we did not miss any study that reported relevant subgroup results. In the actual meta-analyses of CV, we only included data for aNSCLC patients. Some of these studies included both treatment-naïve and treatment-experienced patients, however, we do not have reason to believe that this has (externally) biased the estimates of diagnostic accuracy focused on the presence of driver mutation for the 1L target population. If we excluded studies comprising not only treatment-naïve patients, the evidence base would have been limited.

The available evidence base regarding the CV and CU of ctDNA was limited. The CV studies had small sample sizes which made it challenging to reliably estimate sensitivity for rare driver mutations. Regarding the risk of bias, for only four studies we did not identify any concerns. Although we did not observe a difference in overall diagnostic performance between higher and lower quality studies, the limited evidence base makes interpretation of this observation difficult. Similarly, the limited number of studies did not make it feasible to reliably evaluate potential drivers of between-study differences in ctDNA testing performance either.

Despite the limitations of the available evidence base, the CV findings from our study can have important clinical implications for the role of ctDNA testing in NSCLC. Our results support current guidelines that recommend ctDNA testing as a complementary approach rather than a replacement for TB to inform 1L therapy, unless there is insufficient tissue for NGS or risks of biopsy are excessive.^7,65^ This is particularly relevant for specific driver mutations such as *EGFR* and *KRAS*, which had much higher detection rates than *ROS1* or *ALK*. While multiple factors have been implicated in the sensitivity of ctDNA including tumor burden and the presence of osseous metastases^68^, molecular heterogeneity has been relatively unexplored as a predictor of sensitivity for ctDNA. In particular, molecular fusions such as *ALK* and *ROS1* may be less detectable via ctDNA given the number of potential fusion partners which makes development of a sensitive ctDNA assay challenging.^66,67^ For example, in a patient who has never smoked where risks of biopsy may be high or the initial biopsy may not have sufficient tissue, a negative ctDNA test may not be sufficient to rule out the presence of a *ROS1* fusion which has profound implications for clinical decision-making around repeat biopsies at time of diagnosis and in the future. In addition to informing 1L therapy, ctDNA findings before initiating treatment will also help with the interpretation of ctDNA to monitor disease progression and tumor burden. For example, the absence of a mutation at follow-up that was detected upfront is indicative of effective treatment.^69^ Ongoing presence of a particular mutation after initiation of therapy (even in the absence of a targetable mutation) may reflect a higher underlying tumor burden and an independent risk factor for survival.^70^

There are only two studies that have evaluated the impact of the CU of ctDNA in terms of PFS in 1L aNSCLC patients, and no studies evaluating OS. This is a key area of future study, as baseline ctDNA may rarely pick up a mutation that would not be detected with tissue biopsy and influence treatment decisions, ultimately impacting OS. Additionally, we only included PFS and OS in our assessment of CU of ctDNA, and future studies may also want to examine other CU measures such as turn-around time of test results, time to initiation of therapy, number of patients matched to targeted therapy, or treatment decision impacts.

## Conclusion

Based on the currently available evidence, ctDNA testing in patients with aNSCLC has an overall acceptable diagnostic accuracy for detecting any guideline-recommended oncogenic driver mutation when using TB as the reference standard. However, the sensitivity of ctDNA testing varies greatly depending on the specific oncogenic driver mutation. At the time of this review, there were limited studies on less common driver mutations, and this is an essential area of future research. Given the current detection rates, ctDNA cannot be recommended as a replacement for TB, considering the actionable driver mutations of interest, unless there is insufficient tissue for TB or risks associated with the procedure. As the technologies around ctDNA testing and NGS analysis continue to evolve, we anticipate new studies will become available at a rapid pace, necessitating timely updates to this systematic review and meta-analysis.

## Data Availability

All data produced in the present study are available upon reasonable request to the authors

## Author Contributions

*Concept and design:* Jansen, Chen.

*Acquisition, analysis, or interpretation of data*: All authors.

*Drafting of the manuscript*: Chen, Jansen, Ragavan

*Critical revision of the manuscript for important intellectual content*: All authors.

*Statistical analysis*: Chen, Jansen

*Administrative, technical, or material support*: Douglas, Phillips

*Supervision*: Jansen, Phillips

## Disclaimer

The National Human Genome Research Institute and the National Cancer Institute had no role in the preparation, review, or approval of the manuscript or decision to submit the manuscript for publication.

