## Supplemental Data for "Clinical validity and utility of circulating tumor DNA (ctDNA) testing in advanced non-small cell lung cancer (aNSCLC): a systematic literature review and meta-analysis"

Table S1: Study eligibility criteria

| Criteria | Description |
| --- | --- |
| **Population** | - Adult advanced (stage III/IV) NCSLC Cancer Patients of any/all race/ethnicity with any/all of the following genomic alterations: - *EGFR* - *ALK* - *ROS1* - *BRAF* - *NTRK1/2/3* - *RET* - *MET* - PD-L1 - *KRAS* - Wildtype - Unknown driver mutation |
| **Interventions** | Any of the following technologies for detection of targetable genomic alterations:   - ctDNA-based genomic profiling - Tissue-based genomic profiling   AND, when outcome of interest is clinical utility:   - At least 80% of population receiving Standard of Care by driver mutation (according to current NCCN Guidelines, table on page 92 of version 3.2022) defined as:   - *EGFR* Exon 19 deletion or L858R: afatinib or erlotinib or dacomitinib or gefitinib or osimertinib or ramucirumab or bevacizumab;   - *EGFR* S768I or L861Q, and/or G719X: afatinib, or erlotinib, or dacomitinib, or gefitinib, or osimertinib;   - *EGFR* Exon 20 insertion mutation positive: amivantamab-vmjw or mobocertinib;   - *ALK* rearrangement positive: alectinib or brigatinib or certinib or crizotinib or lorlatinib   - *ROS1* rearrangement positive: certinib or crizotinib or entrectinib   - *BRAF* V600E mutation positive: dabrafenib/trametinib or vemurafenib   - *NTRK*1/2/3: Larotrectinib or entrectinib   - *RET*: selpercatinib or pralsetinib or cabozantinib   - *MET*: capmatinib or crizotinib or tepotinib   - *KRAS*: sotorasib   - PD-L1: pembrolizumab or carboplatin or cisplatin or pemetrexed or paclitaxel or bevacizumab or atezolizumab or albumin-bound paclitaxel or nivolumab or ipilimumab   - Wildtype:   [NOTE: Studies must contain two or more driver genes (e.g., EGFR and ALK) in order to be included] |
| **Outcomes** | - Clinical validity   - Sensitivity;   - Specificity; or   - Positive/negative predictive value   OR   - Clinical utility   - Progression free survival; or   - Overall survival, reported as KM or survival curve |
| **Study design** | - Prospective or retrospective cohort studies - Randomized controlled trials |
| **Language** | - English |
| **Time** | - Only studies published from year 2012 onward |

Table S2: Search strategy with Medline, Embase - searched within Embase platform (on 7/5/2023)

| **No.** | **Criteria** | **Strings** | **Hits** |
| --- | --- | --- | --- |
| 1 | Population | 'non small cell lung cancer'/exp OR 'non small cell lung cancer' OR nsclc | 226949 |
| 2 | Population | egfr OR 'epidermal growth factor receptor' | 275762 |
| 3 | Population | alk OR 'anaplastic lymphoma receptor tyrosine kinase' | 27898 |
| 4 | Population | ros1 OR 'ros proto-oncogene 1 receptor tyrosine kinase' | 5689 |
| 5 | Population | braf OR 'b-raf proto-oncogene, serine/threonine kinase' | 43312 |
| 6 | Population | ntrk1 OR 'neurotrophic receptor tyrosine kinase 1' | 2063 |
| 7 | Population | ntrk2 OR 'neurotrophic receptor tyrosine kinase 2' | 1162 |
| 8 | Population | ntrk3 OR 'neurotrophic receptor tyrosine kinase 3' | 1762 |
| 9 | Population | ret OR 'ret proto-oncogene' | 18947 |
| 10 | Population | met OR 'met proto-oncogene, receptor tyrosine kinase' | 420609 |
| 11 | Population | kras OR 'kras proto-oncogene' | 49111 |
| 12 | Population | 'pd-l1' OR 'programmed death-ligand 1' | 51488 |
| 13 | Population | 'genomic alterat*' | 12040 |
| 14 | Population | wildtype | 355241 |
| 15 | Population | #2 OR #3 OR #4 OR #5 OR #6 OR #7 OR #8 OR #9 OR #10 OR #11 OR #12 OR #13 OR #14 | 1162656 |
| 16 | Population | #1 AND #15 | 71563 |
| 17 | Intervention | 'genomic profiling' OR 'genetic profiling' OR 'molecular profil*' OR 'tumor profil*' OR 'mutation profil*' OR 'molecular test' OR 'biomarker test*' OR 'tumor sequenc*' | 38880 |
| 18 | Intervention | 'liquid biopsy' OR 'fluid biopsy' OR 'cell free dna' OR cfdna OR 'circulating tumor dna' OR 'ctdna' | 34466 |
| 19 | Intervention | 'tissue biopsy' OR 'needle biopsy' | 67521 |
| 20 | Intervention | #17 OR #18 OR #19 | 136618 |
| 21 | Outcomes | 'clinical validity' | 3332 |
| 22 | Outcomes | sensitivity' | 1800575 |
| 23 | Outcomes | 'specificity' | 1078876 |
| 24 | Outcomes | 'positive predictive value' | 77283 |
| 25 | Outcomes | 'negative predictive value' | 68402 |
| 26 | Outcomes | #21 OR #22 OR #23 OR #24 OR #25 | 2264504 |
| 27 | Outcomes | 'clinical utility' | 50221 |
| 28 | Outcomes | 'progression free survival' OR 'pfs' | 206013 |
| 29 | Outcomes | 'overall survival' OR 'os' | 627143 |
| 30 | Outcomes | #27 OR #28 OR #29 | 733394 |
| 31 | Outcomes | #26 OR #30 | 2945799 |
| 32 | Study design | 'retrospective study' OR 'prospective study' OR 'observational study' OR 'cohort analysis' OR 'real world evidence' OR 'register' OR 'electronic health record' OR 'electronic medical record' | 3307459 |
| 33 | Study design | 'clinical trial' OR 'randomized controlled trial' OR 'controlled clinical trial' | 2259219 |
| 34 | Study design | 'single arm trial' | 1825 |
| 35 | Study design | #32 OR #33 OR #34 | 5169692 |
| 36 |  | #16 AND #20 AND #31 AND #35 | 1920 |
| 37 | Limits | #36 AND [english]/lim | 1916 |
| 38 | Limits | #37 AND (2012:py OR 2013:py OR 2014:py OR 2015:py OR 2016:py OR 2017:py OR 2018:py OR 2019:py OR 2020:py OR 2021:py OR 2022:py) | 1749 |

Table S3. Patient characteristics in included clinical validity (CV) studies (n=18)

|  | **No. of Patients** | **Tumor stage III/IV (advanced)** | **Age (years)** | **Male** | **Former/Current Smokers** | **Caucasian** | **Asian** | **African American** | **Hispanic** |
| --- | --- | --- | --- | --- | --- | --- | --- | --- | --- |
| **Author_Year** | n | % | Median (range) | n (%) | n (%) | n (%) | n (%) | n (%) | n (%) |
| Bustamante Alvarez_2021 | 94 | 100 | NA | NA | NA | NA | NA | NA | NA |
| Fernandes_2021 | 115 | 100 | 66 (38-92) | 71 (61.7%) | 73 (63.5%) | NA | NA | NA | NA |
| Leighl_2019 | 282 | 100 | 69 (26-100) | 129 (45.7%) | 214 (76.1%) | 231 (81.9%) | 17 (6%) | 18 (6.4%) | 23 (8.2%) |
| Li_2019 | 127 | 100 | NA | 47 (37%) | NA | NA | NA | NA | NA |
| Lin_2021 | 100 | 98 | 67.5 (35-92) | 44 (44%) | NA | NA | NA | NA | NA |
| Müller_2017 | 98 | 54.1 | mean=66 | 43 (44%) | NA | NA | NA | NA | NA |
| Palmero_2021 | 186 | 100 | 63.4 (39-86) | 121 (65%) | 132 (71%) | 180 (97%) | 0 (0%) | 0 (0%) | 0 (0%) |
| Park_2021 | 262 | 100 | 62 (30-89) | 175 (66.8%) | 155 (59.2%) | NA | NA | NA | NA |
| Pritchett_2019 | 264 | 100 | mean=67.1 (SD=11) | 128 (48.5%) | 225 (85.2%) | 226 (85.6%) | 6 (2.3%) | 26 (9.8%) | 0 (0%) |
| Schwaederlé_2017 | 88 | 100 | 66.2 (36.3-89.5) | 30 (34%) | 50 (56.8%) | 58 (66%) | 18 (20.5%) | 1 (1.1%) | 2 (2.3%) |
| Yao_2017 | 39 | 100 | 62 (28-78) | 19 (48.7%) | 10 (25.6%) | NA | NA | NA | NA |
| Xu_2016 | 42 | 100 | 62 (37-75) | 21 (50%) | NA | NA | NA | NA | NA |
| Liu_2018 | 72 | 100 | 59 (40-83) | 44 (61.1%) | 29 (40.3%) | NA | NA | NA | NA |
| Couraud_2014 | 106 | 87.5 | Mean=68.7 (SD=13.7) | 13 (12.3%) | 0 | NA | NA | NA | NA |
| Chen_2019 | 50 | 40 | NA | 35 (70%) | 21 (42%) | NA | NA | NA | NA |
| Remon_2019 | 214 | 100 | Mean=63.7 (SD=10.53) | 126 (58.9%) | 175 (83.3%) | NA | NA | NA | NA |
| Guo_2016 | 41 | 26.8 (IV) | 52 (38-73) | 22 (53.7%) | NA | NA | NA | NA | NA |
| Sabari_2019 | 210 | 100 | 65 (28-91) | 86 (41%) | NA | NA | NA | NA | NA |

Table S4. Data included in meta-analysis on the clinical validity (CV) of ctDNA testing for multi-gene detection (n=13)

| **Author_Year** | **No. of Patients** | **Test driver genes** | **TP** | **FP** | **FN** | **TN** | **Total No. of Samples** |
| --- | --- | --- | --- | --- | --- | --- | --- |
| Bustamante Alvarez_2021 | 94 | Overall (*EGFR*, *ALK* fusions, *BRAF*, *MET* exon 14 skipping mutation, *ROS1* mutations and *RET* fusions) | 22 | 7 | 11 | 54 | 94 |
| Fernandes_2021 | 127 | Overall (Oncomine – *EGFR, KRAS, ALK, BRAF, MET, ROS1, ERBB2, MAP2K1, NRAS, PIK3CA*, and *TP53*) | 51 | 3 | 12 | 61 | 127 |
| Leighl_2019 | 282 | Overall (*EGFR, ALK* fusion, *BRAF* V600E, *MET* amplification, *MET* exon 14 skipping, *ROS1* fusion, *RET* fusion, *ERBB2* mutation) | 35 | 0 | 9 | 862 | 906 |
| Li_2019 | 91 for sensitivity analysis; 19 for specificity analysis | Overall (*EGFR, KRAS, ALK, BRAF, MET, ROS1, RET, ERBB2*) | 68 | 0 | 23 | 789 | 880 |
| Müller_2017 | 29 | Key 6 genes (*EGFR, KRAS, ALK, BRAF, ROS1, TP53*) | 16 | 1 | 12 | 145 | 174 |
| Park_2021 | 262 | Key genes (*EGFR* Del 9, *EGFR* L858R, *KRAS*, *ALK*, *BRAF* V600E, *MET* exon 14, *MET* amplification, *ROS1*, *RET*, *ERBB2* exon 20) | 95 | 19 | 69 | 2437 | 2620 |
| Pritchett_2019 | 95 | Key 8 genes (*EGFR* (exons 18-21), *KRAS*, *ALK/ROS1*, *BRAF* V600E, *MET* exon 14, *ERBB2* exon 20, *STK11*) | 88 | 2 | 31 | 1027 | 1148 |
| Yao_2017 | 39 | Overall (EGFR, KRAS, PIK3CA, ALK and gene fusions (EML4-ALK and KIF5B-RET)) | 19 | 0 | 9 | 128 | 156 |
| Xu_2015 | 42 | Key genes (*EGFR, KRAS, BRAF, ERBB2, PIK3CA, TP53*) | 23 | 25 | 7 | 197 | 252 |
| Liu_2017 | 72 | Overall (*EGFR* L858R, *EGFR* exon 19 del, *KRAS* G12X, *ALK* rearrangement) | 34 | 1 | 16 | 222 | 273 |
| Couraud_2014 | 50 | All 12 amplicons together (*EGFR* (exons 18-21), *KRAS* (exons 2 & 3), *BRAF* (exons 11 & 15), *ERBB2* (exons 19 & 20), *PI3KCA* (exons 9 & 20)) | 24 | 1 | 22 | 489 | 536 |
| Remon_2019 | 94 | Core gene variants (*EGFR* (exons 18-21), *KRAS, BRAF* V600E, *MET* exon 14, *ERBB2* exon 20, *STK11*) | 35 | 10 | 8 | 320 | 373 |
| Sabari_2019 | 106 | *Key genes (EGFR, KRAS, ALK, BRAF, MET, ROS1, RET)* | 44 | 7 | 35 | 656 | 742 |

TP: true positive; FP: false positive; FN: false negative; TN: true negative

Figure S1. Bayesian Posterior Predictive Contours (50%, 75%, 95%) of ctDNA testing for multi-gene detection (n=13)

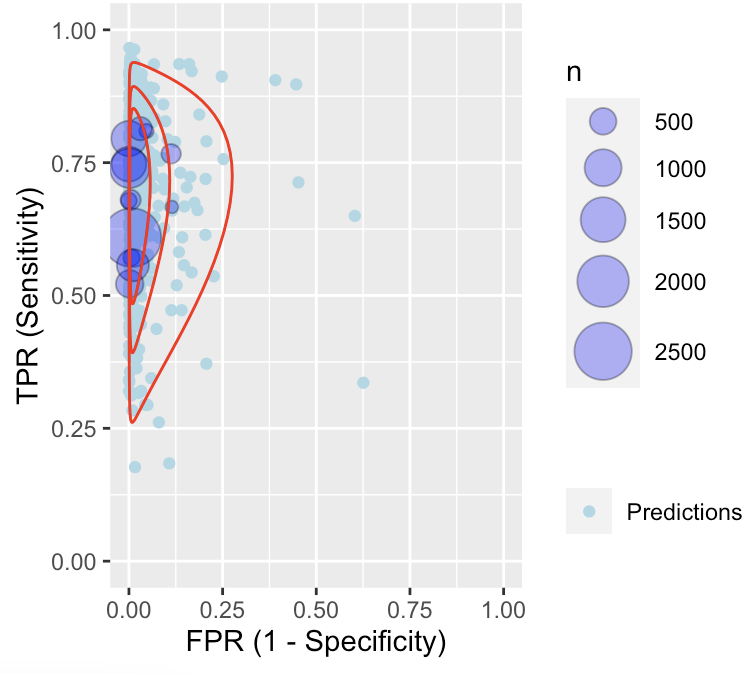

Each purple circle identifies the true positive rate (TPR) vs. the false positive rate (FPR) of each study (observed data). Light blue circles are predictions based on Bayesian meta-analysis of data by fitting a bivariate random effects model, with red lines indicating parametric predictive contours.

Table S5. Results from frequentist and Bayesian bivariate random effects meta-analyses

|  |  | Frequentist Approach | | Bayesian Approach | | |
| --- | --- | --- | --- | --- | --- | --- |
|  | No. of studies | Sensitivity (95% CI) | Specificity (95% CI) | Sensitivity (95% CI) | Specificity (95% CI) | BAUC |
| Overall | 13 | 0.69(0.63, 0.74) | 0.99 (0.97, 1.00) | 0.70 (0.60, 0.79) | 0.99 (0.97, 1.00) | 0.71 (0.68, 0.73) |
| *EGFR* | 14 | 0.68 (0.62, 0.74) | 0.98 (0.95, 0.99) | 0.70 (0.60, 0.78) | 0.98 (0.95, 0.99) | 0.71 (0.68, 0.73) |
| *KRAS* | 11 | 0.77 (0.63, 0.86) | 0.96 (0.90, 0.98) | 0.82 (0.70, 0.90) | 0.97 (0.93, 0.99) | 0.72 (0.69, 0.73) |
| *BRAF* | 8 | 0.60 (0.39, 0.77) | 0.99 (0.99, 1.00) | 0.64 (0.37, 0.86) | 1.00 (0.99, 1.00) | 0.53 (0.51, 0.54) |
| *ALK* | 6 | 0.59 (0.45, 0.73) | 0.99 (0.98, 1.00) | 0.60 (0.38, 0.79) | 0.99 (0.98, 1.00) | 0.62 (0.58, 0.64) |
| *ROS1* | 4 | 0.29 (0.13, 0.53) | 0.99 (0.97, 1.00) | 0.26 (0.06, 0.58) | 0.99 (0.97, 1.00) | 0.67 (0.52, 0.73) |
| *MET* | 5 | 0.47 (0.33, 0.62) | 0.98 (0.97, 0.99) | 0.48 (0.26, 0.71) | 0.99 (0.97, 1.00) | 0.70 (0.63, 0.73) |
| *RET* | 3 | 0.38 (0.21, 0.58) | 0.99 (0.97, 1.00) | 0.37 (0.11, 0.71) | 0.98 (0.95, 1.00) | 0.66 (0.50, 0.73) |

Table S6. Data included in meta-analysis on the clinical validity (CV) of ctDNA detection of *EGFR* (n=14)

| **Author_Year** | **No. of patients** | **TP** | **FP** | **FN** | **TN** |
| --- | --- | --- | --- | --- | --- |
| Leighl_2019 | 223 | 27 | 0 | 5 | 414 |
| Li_2019 | 91 for sensitivity analysis, 19 for specificity analysis | 29 | 0 | 8 | 73 |
| Müller_2017 | 29 | 6 | 0 | 1 | 22 |
| Palmero_2021 | 148 | 24 | 0 | 12 | 112 |
| Park_2021 | 262 | 35 | 4 | 14 | 471 |
| Pritchett_2019 | 164 | 13 | 0 | 5 | 146 |
| Schwaederlé_2017 | 26 | 7 | 2 | 3 | 14 |
| Yao_2017 | 39 | 12 | 0 | 5 | 22 |
| Xu_2015 | 42 | 4 | 11 | 3 | 24 |
| Liu_2017 | 72 | 26 | 0 | 12 | 106 |
| Couraud_2014 | 45 | 19 | 1 | 8 | 147 |
| Chen_2019 | 7 | 2 | 0 | 1 | 4 |
| Remon_2019 | 94 | 7 | 1 | 1 | 78 |
| Sabari_2019 | 106 | 24 | 1 | 20 | 61 |

TP: true positive; FP: false positive; FN: false negative; TN: true negative

Figure S2. Forest plot of sensitivity and specificity on ctDNA detection of *EGFR* from bivariate random effects meta-analyses (n=14)

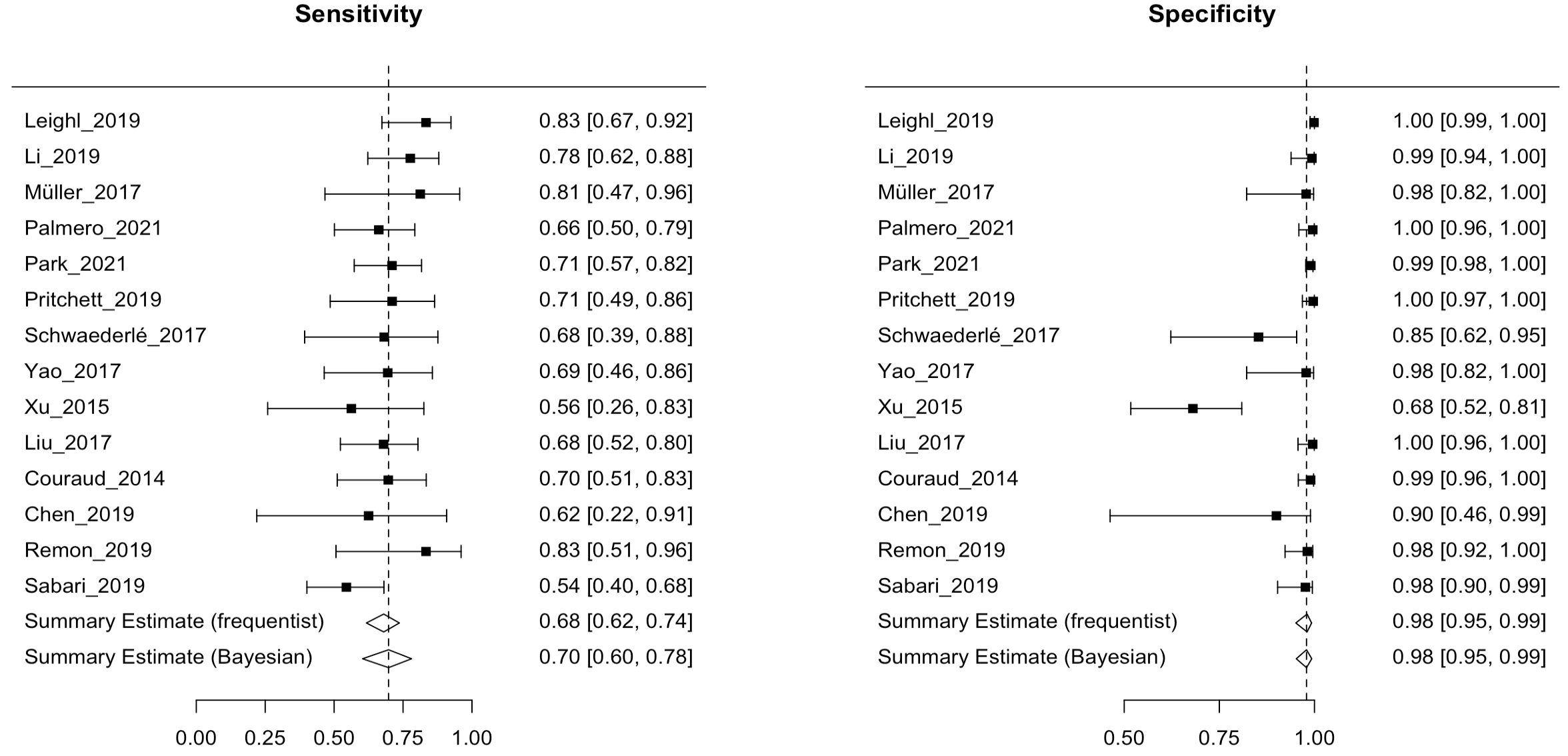

Sensitivity and specificity of single studies were based on frequentist estimations; I^2^ = 0% based on the frequentist approach

Figure S3. Summary receiver operating characteristics (SROC) plots based on bivariate random-effects meta-analyses of ctDNA detection of *EGFR* (n=14)

(a) SROC (frequentist approach):

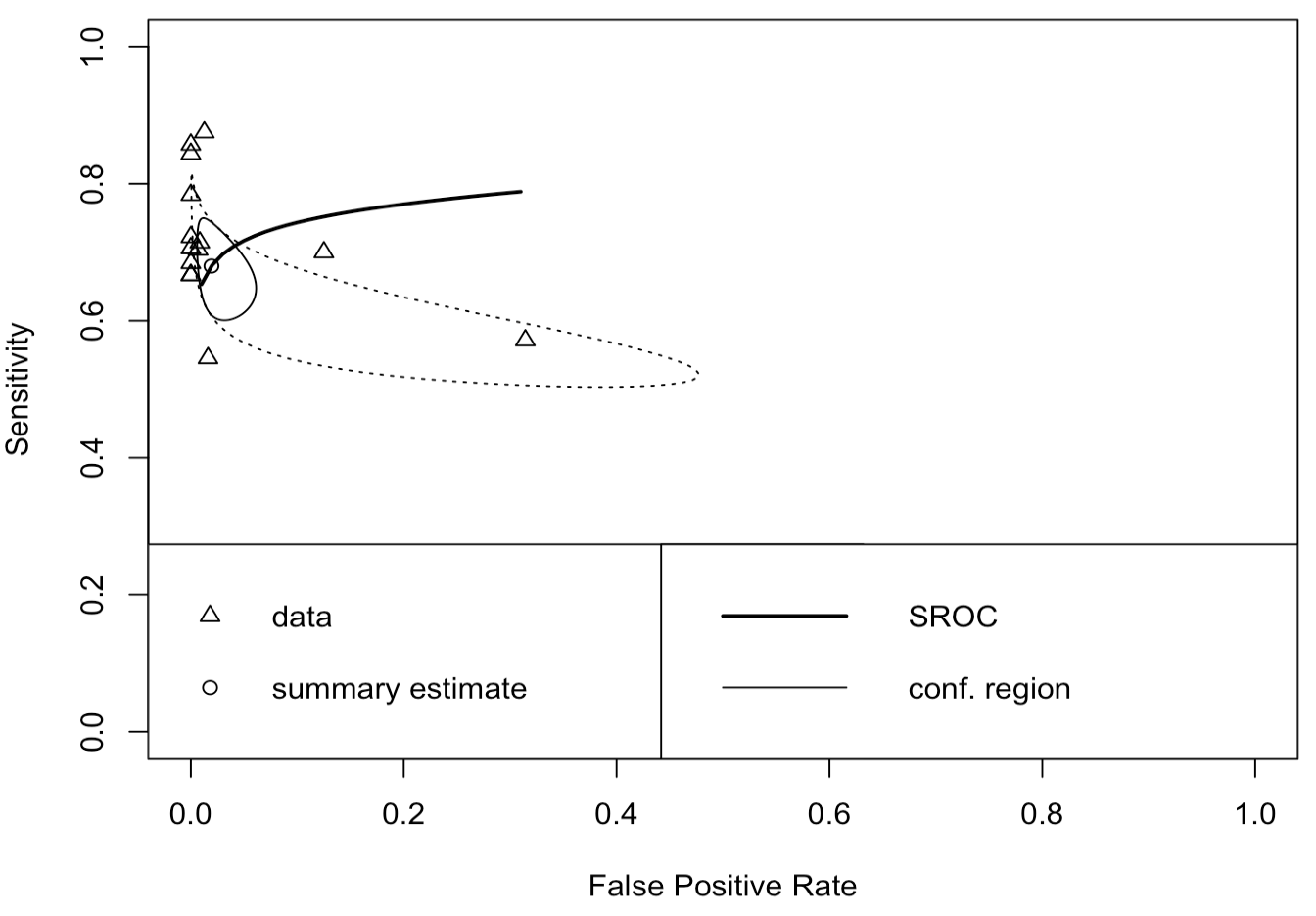

(b) SROC (Bayesian approach):

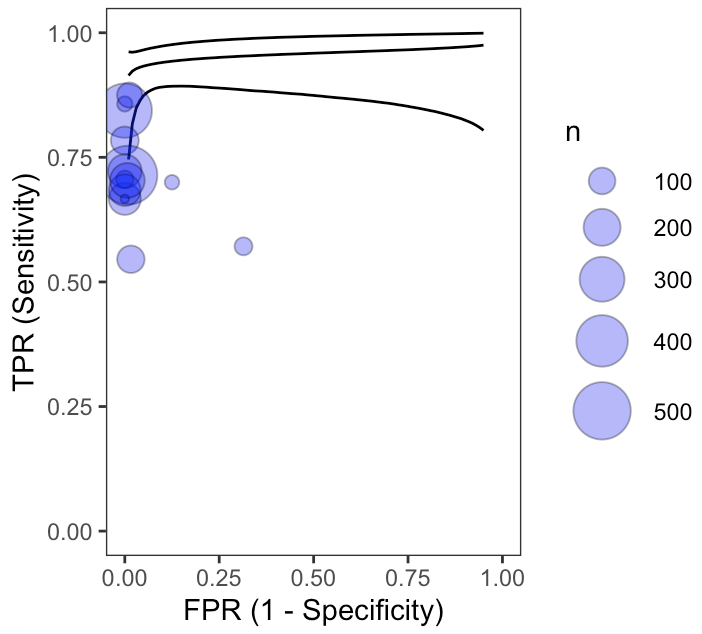

Figure S4. Bayesian Posterior Predictive Contours (50%, 75%, 95%) for ctDNA detection of *EGFR* (n=14)

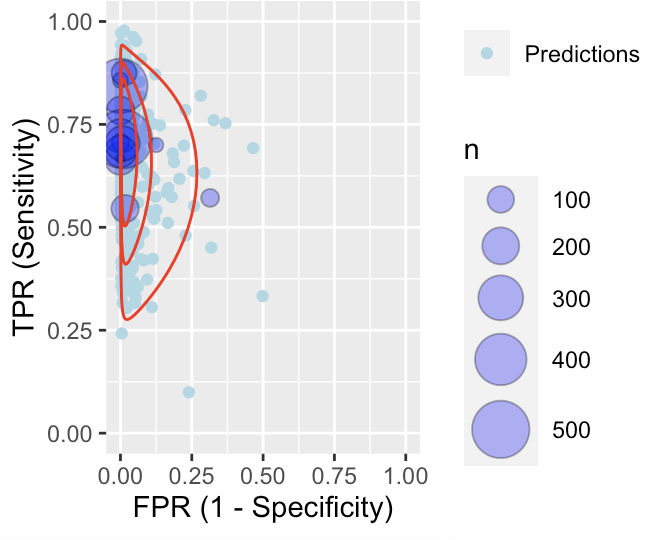

Each purple circle identifies the true positive rate (TPR) vs. the false positive rate (FPR) of each study (observed data). Light blue circles are predictions based on Bayesian meta-analysis of data by fitting a bivariate random effects model, with red lines indicating parametric predictive contours.

Table S7. Data included in meta-analysis on the CV of ctDNA detection of *KRAS* (n=11)

| **Author_Year** | **No. of patients** | **TP** | **FP** | **FN** | **TN** |
| --- | --- | --- | --- | --- | --- |
| Bustamante Alvarez_2021 | 94 | 69 | 9 | 2 | 14 |
| Li_2019 | 91 for sensitivity analysis, 19 for specificity analysis | 23 | 0 | 6 | 81 |
| Müller_2017 | 29 | 4 | 0 | 5 | 20 |
| Park_2021 | 262 | 23 | 3 | 11 | 225 |
| Pritchett_2019 | 146 | 48 | 1 | 12 | 86 |
| Yao_2017 | 39 | 3 | 0 | 1 | 35 |
| Xu_2015 | 42 | 6 | 6 | 0 | 30 |
| Liu_2017 | 65 | 6 | 0 | 2 | 57 |
| Couraud_2014 | 45 | 2 | 0 | 1 | 82 |
| Remon_2019 | 94 | 22 | 7 | 3 | 56 |
| Sabari_2019 | 106 | 8 | 2 | 10 | 86 |

TP: true positive; FP: false positive; FN: false negative; TN: true negative

Figure S5. Forest plot of sensitivity and specificity on ctDNA detection of *KRAS* from bivariate random effects meta-analyses (n=11)

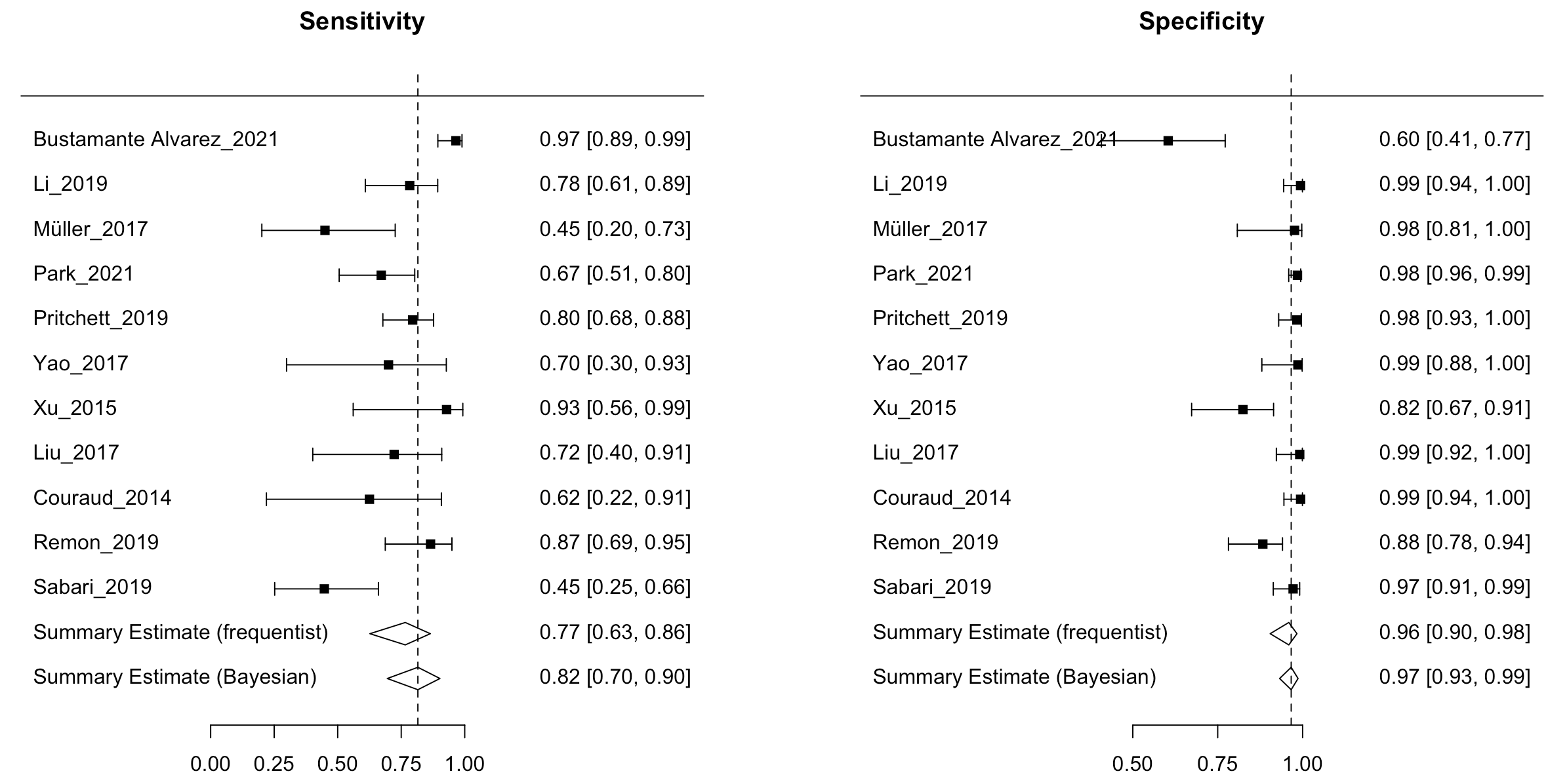

Sensitivity and specificity of single studies were based on frequentist estimations; I^2^ = 0% based on the frequentist approach

Figure S6. Summary receiver operating characteristics (SROC) plots based on bivariate random-effects meta-analyses of ctDNA detection of *KRAS* (n=11)

(a) SROC (frequentist approach)

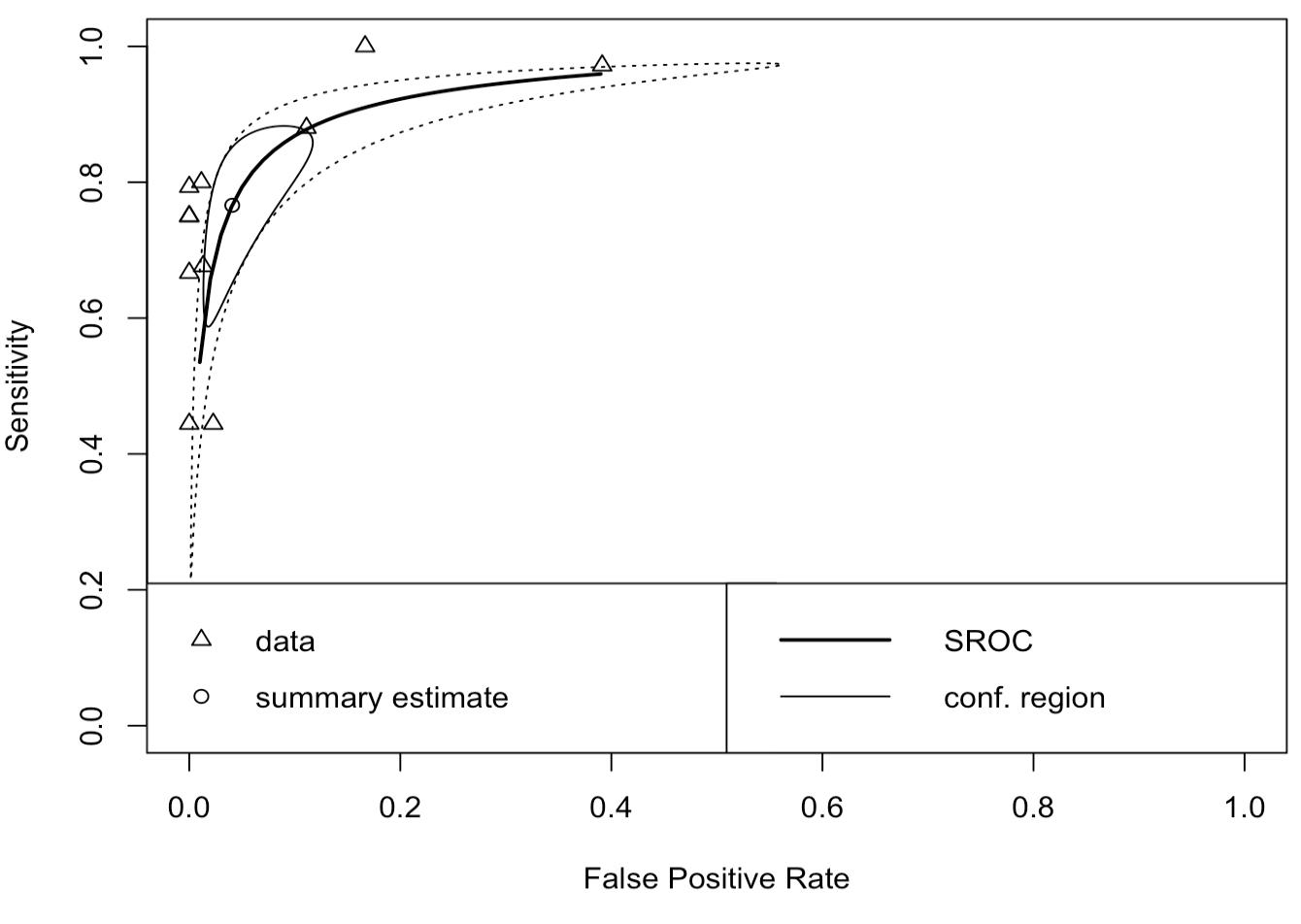

(b) SROC (Bayesian approach)

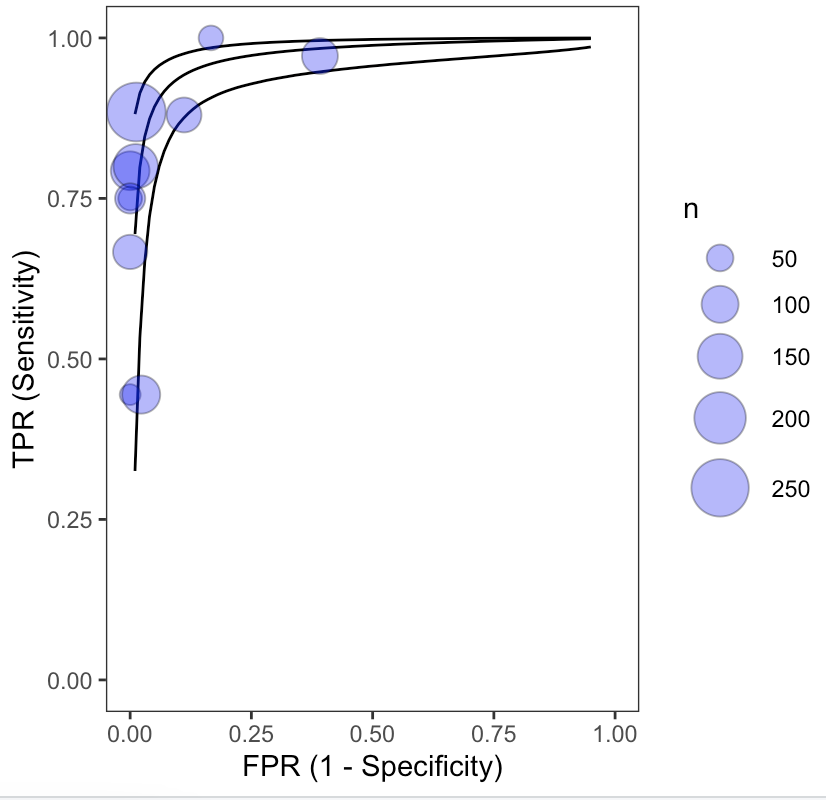

Figure S7. Bayesian Posterior Predictive Contours (50%, 75%, 95%) for ctDNA detection of *KRAS* (n=11)

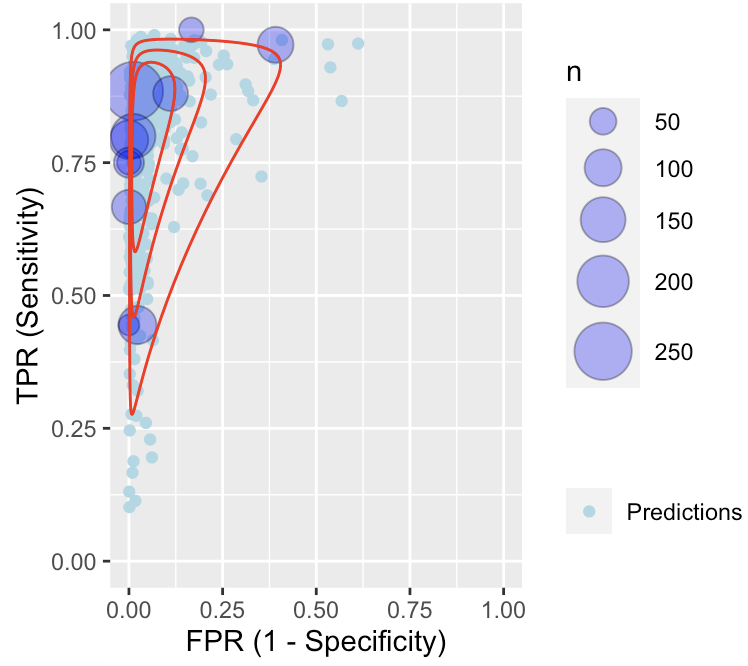

Each purple circle identifies the true positive rate (TPR) vs. the false positive rate (FPR) of each study (observed data). Light blue circles are predictions based on Bayesian meta-analysis of data by fitting a bivariate random effects model, with red lines indicating parametric predictive contours.

Table S8. Data included in meta-analysis on the CV of ctDNA detection of *BRAF* (n=8)

| **Author_Year** | **No. of patients** | **TP** | **FP** | **FN** | **TN** |
| --- | --- | --- | --- | --- | --- |
| Leighl_2019 | 92 | 2 | 0 | 0 | 90 |
| Li_2019 | 91 for sensitivity analysis, 19 for specificity analysis | 3 | 0 | 0 | 107 |
| Müller_2017 | 29 | 0 | 0 | 1 | 28 |
| Park_2021 | 262 | 1 | 1 | 2 | 258 |
| Pritchett_2019 | 151 | 5 | 2 | 0 | 140 |
| Couraud_2014 | 44 | 0 | 0 | 2 | 86 |
| Remon_2019 | 94 | 1 | 0 | 1 | 75 |
| Sabari_2019 | 106 | 3 | 0 | 0 | 103 |

TP: true positive; FP: false positive; FN: false negative; TN: true negative

Figure S8. Forest plot of sensitivity and specificity on ctDNA detection of *BRAF* from bivariate random effects meta-analyses (n=8)

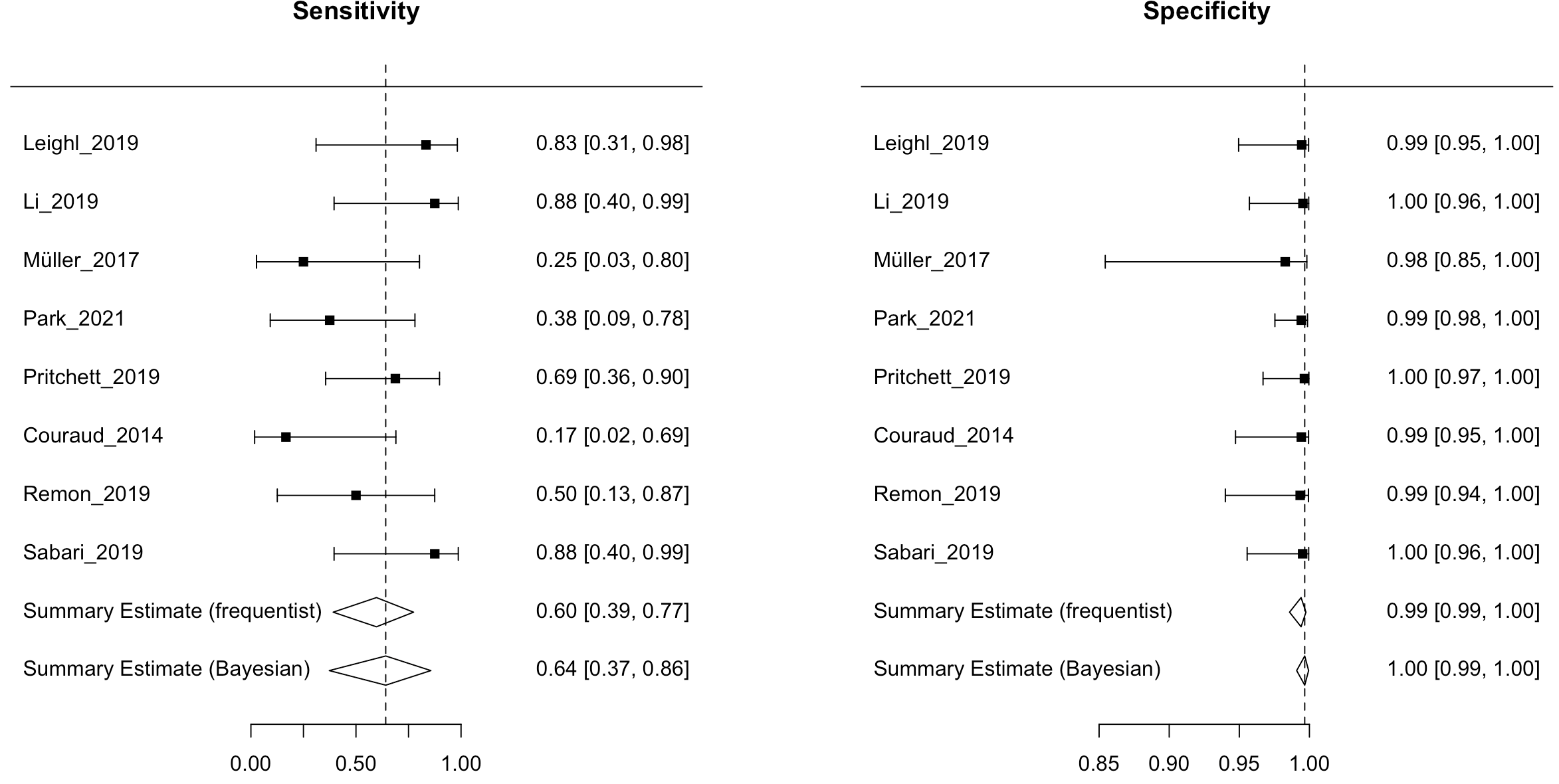

Sensitivity and specificity of single studies were based on frequentist estimations; I^2^ = 0% based on the frequentist approach

Figure S9. Summary receiver operating characteristics (SROC) plots based on bivariate random-effects meta-analyses of ctDNA detection of *BRAF (*n=8)

(a) SROC (frequentist approach):

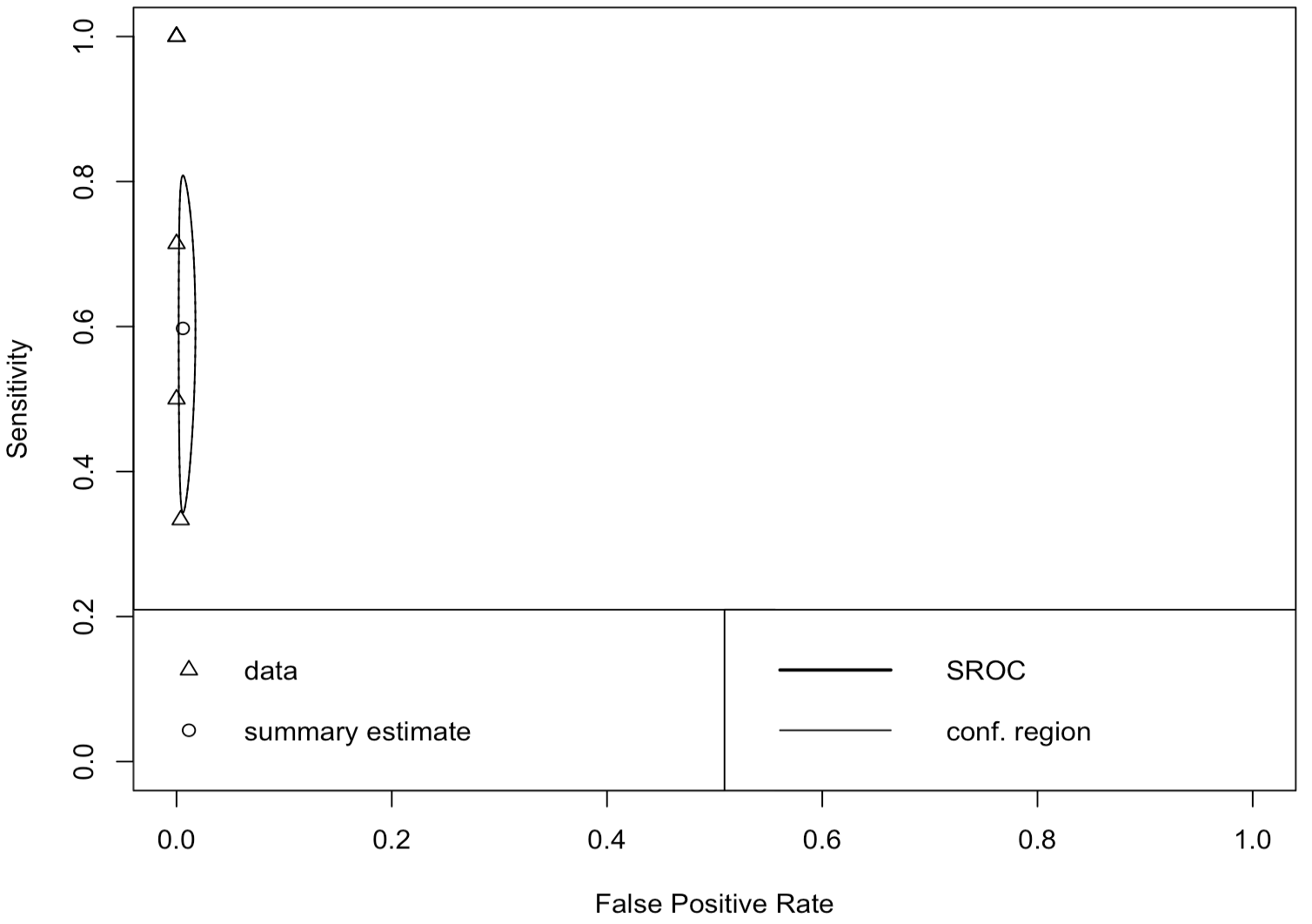

(b) SROC (Bayesian approach):

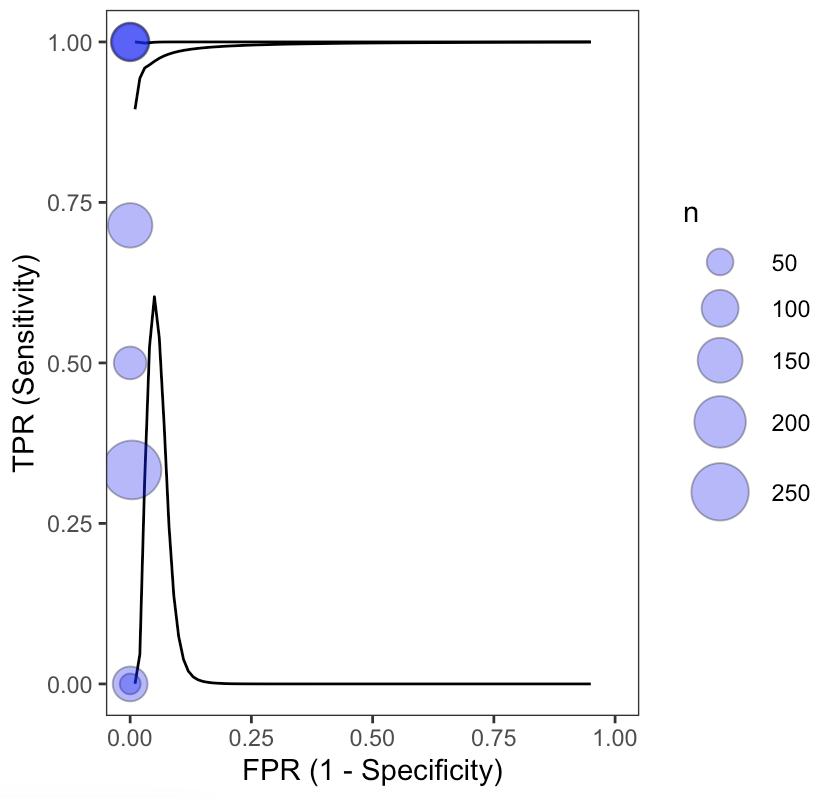

Figure S10. Bayesian Posterior Predictive Contours (50%, 75%, 95%) for ctDNA detection of *BRAF* (n=8)

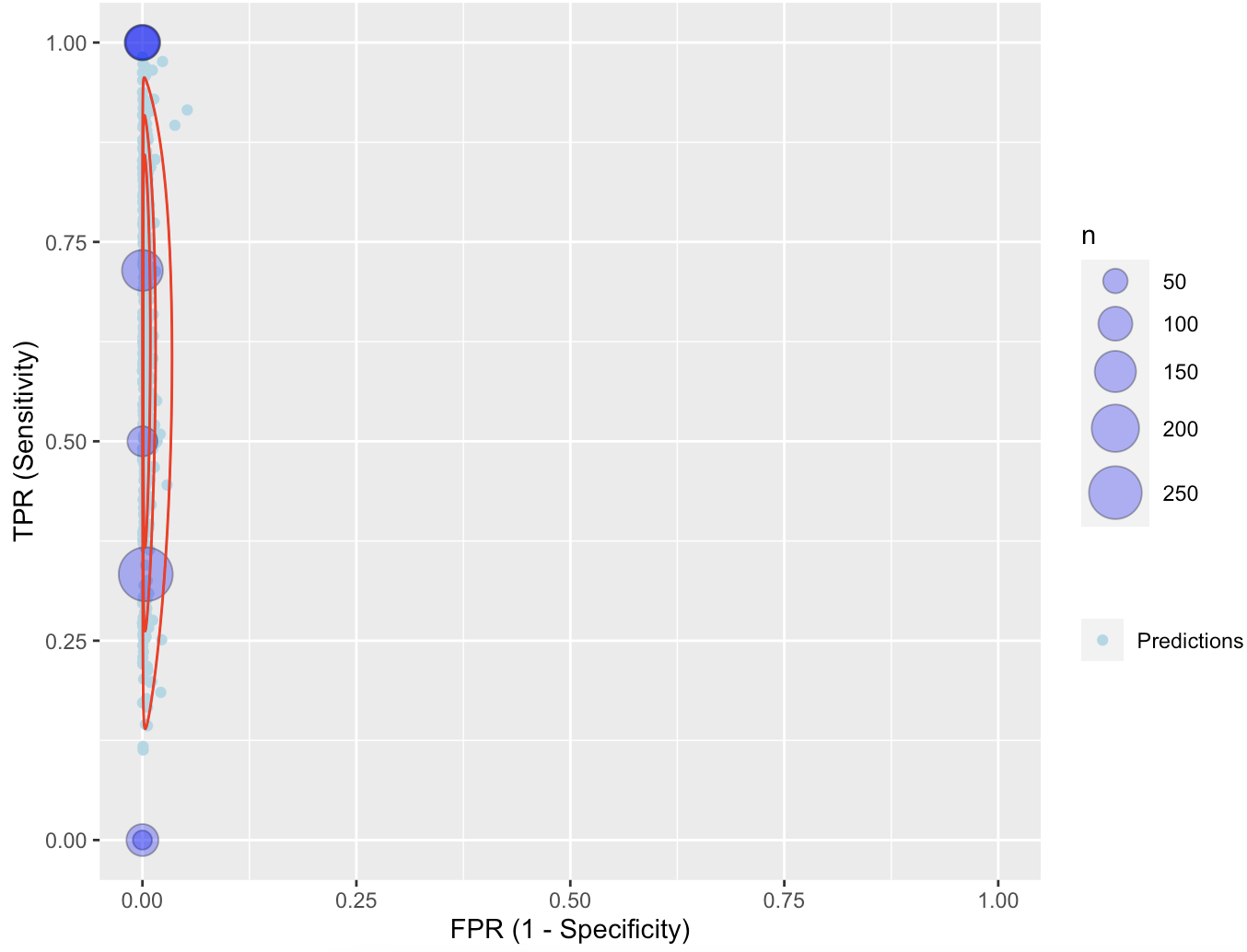

Each purple circle identifies the true positive rate (TPR) vs. the false positive rate (FPR) of each study (observed data). Light blue circles are predictions based on Bayesian meta-analysis of data by fitting a bivariate random effects model, with red lines indicating parametric predictive contours.

Table S9. Data included in meta-analysis on the CV of ctDNA detection of *ALK* (n=6)

| **Author_Year** | **No. of patients** | **TP** | **FP** | **FN** | **TN** |
| --- | --- | --- | --- | --- | --- |
| Leighl_2019 | 215 | 6 | 0 | 2 | 207 |
| Li_2019 | 91 for sensitivity analysis, 19 for specificity analysis | 5 | 0 | 3 | 102 |
| Palmero_2021 | 132 | 2 | 1 | 3 | 126 |
| Park_2021 | 262 | 6 | 0 | 4 | 252 |
| Liu_2017 | 64 | 2 | 1 | 2 | 59 |
| Sabari_2019 | 106 | 4 | 1 | 2 | 99 |

TP: true positive; FP: false positive; FN: false negative; TN: true negative

Figure S11. Forest plot of sensitivity and specificity on ctDNA detection of *ALK* from bivariate random effects meta-analyses (n=6)

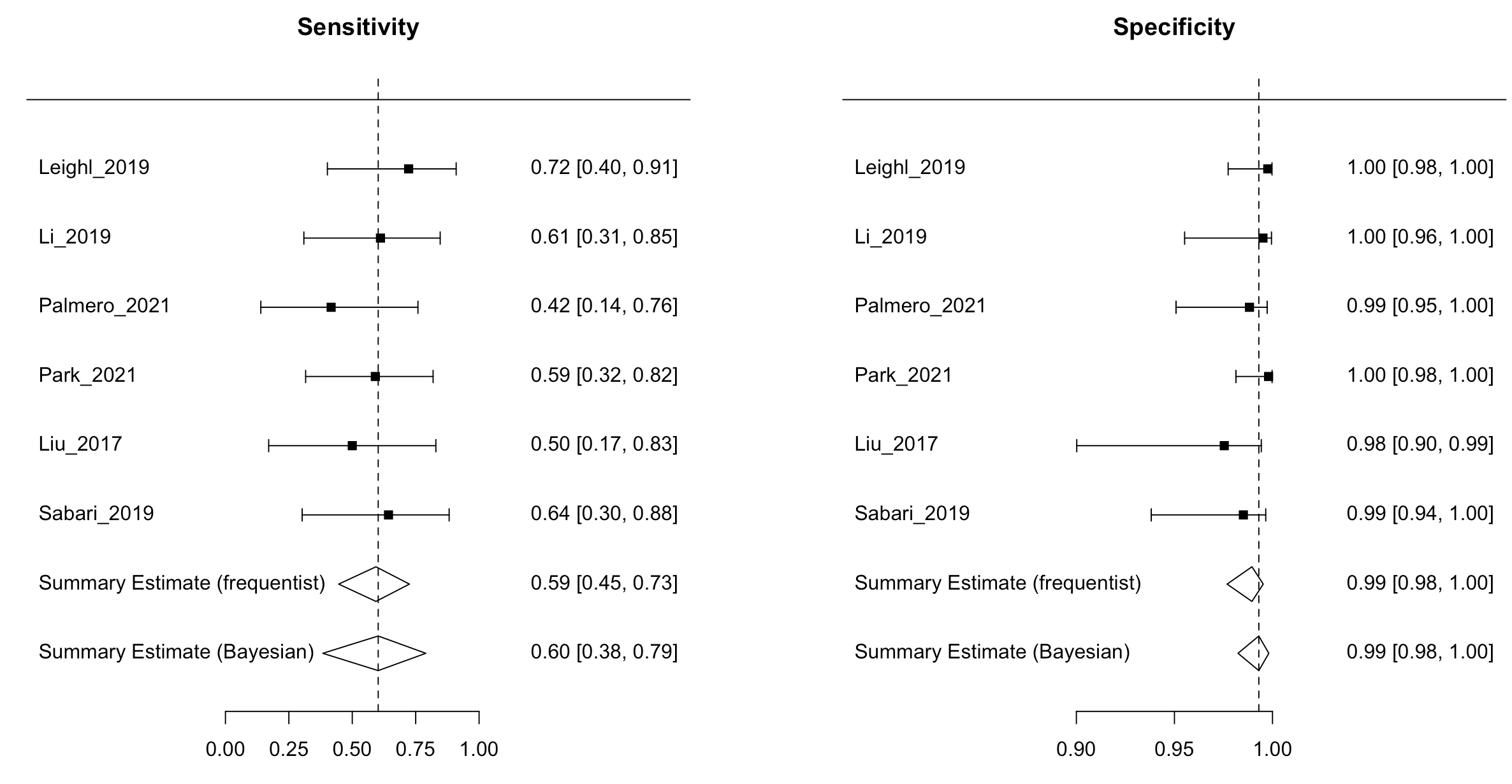

Sensitivity and specificity of single studies were based on frequentist estimations; I^2^ = 10.9% based on the frequentist approach

Figure S12. Summary receiver operating characteristics (SROC) plots based on bivariate random-effects meta-analyses of ctDNA detection of *ALK* (n=6)

(a) SROC (frequentist approach):

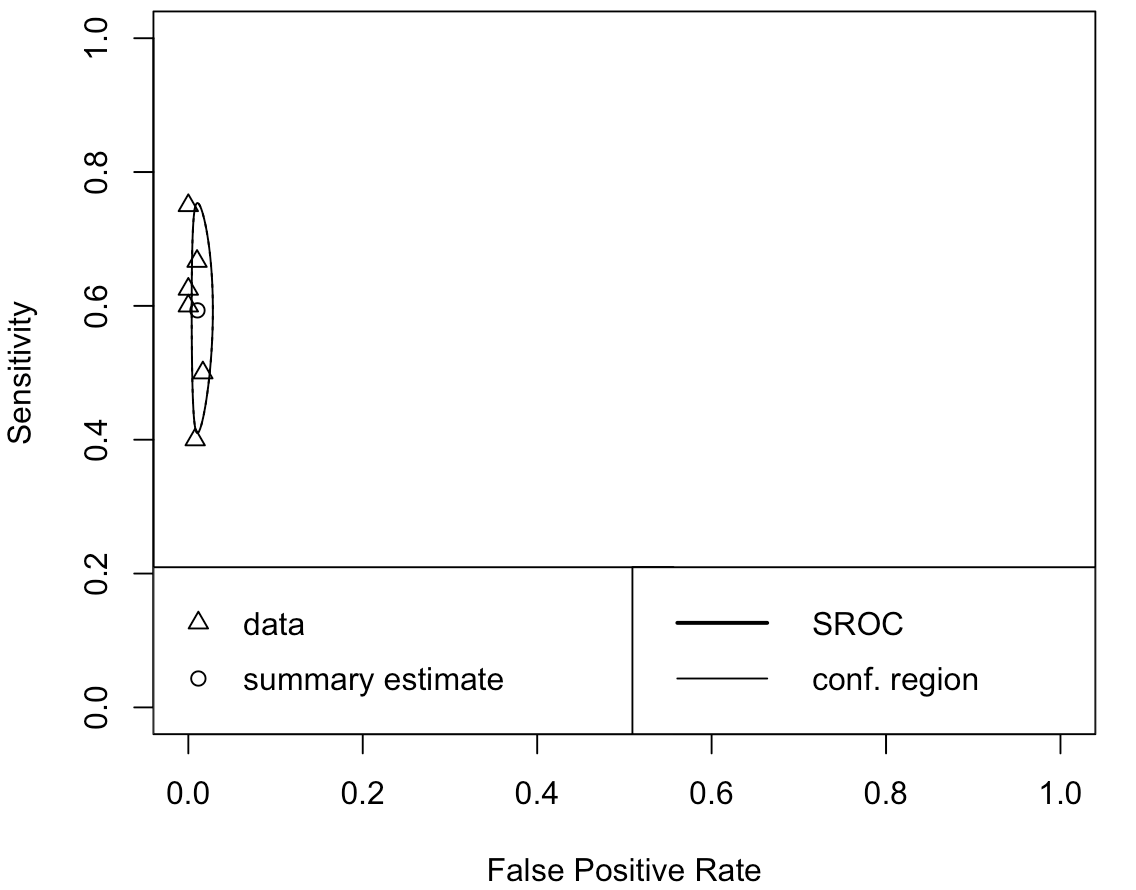

(b) SROC (Bayesian approach):

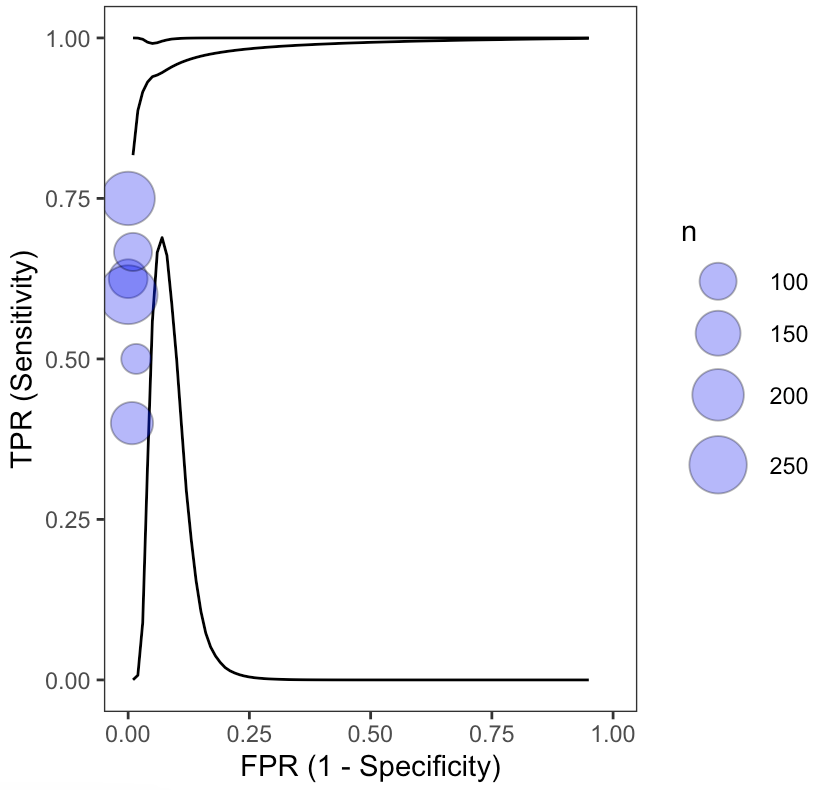

Figure S13. Bayesian Posterior Predictive Contours (50%, 75%, 95%) for ctDNA detection of *ALK* (n=6)

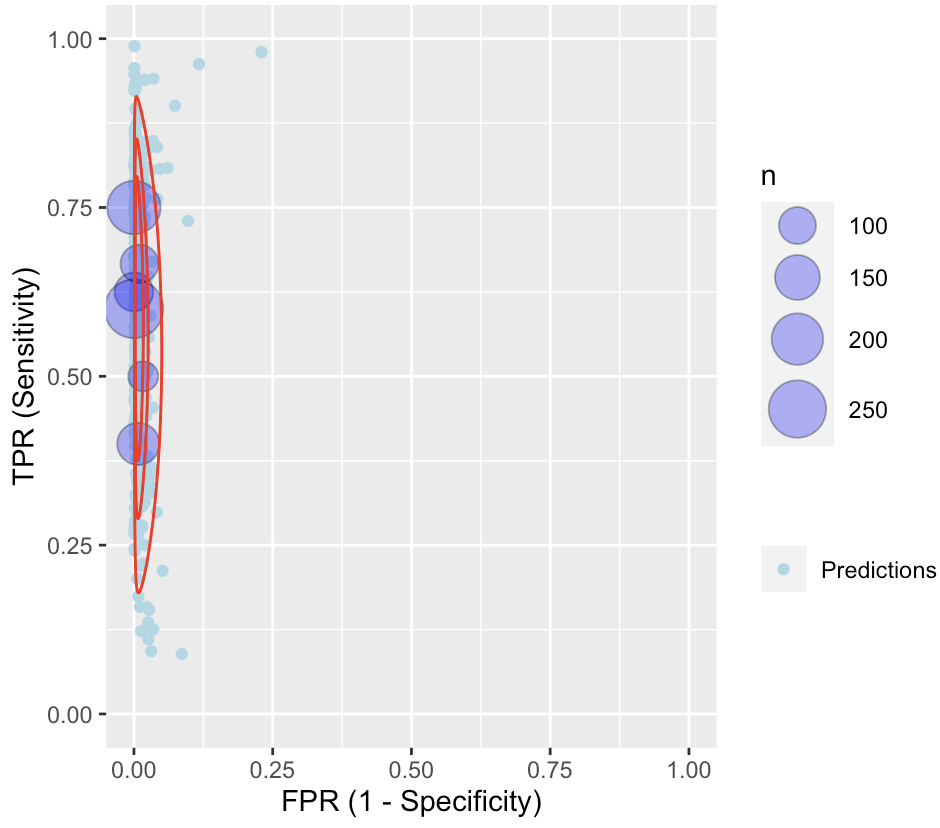

Each purple circle identifies the true positive rate (TPR) vs. the false positive rate (FPR) of each study (observed data). Light blue circles are predictions based on Bayesian meta-analysis of data by fitting a bivariate random effects model, with red lines indicating parametric predictive contours.

Table S10. Data included in meta-analysis on the CV of ctDNA detection of *ROS1* (n=4)

| **Author_Year** | **No. of patients** | **TP** | **FP** | **FN** | **TN** |
| --- | --- | --- | --- | --- | --- |
| Leighl_2019 | 153 | 0 | 0 | 2 | 151 |
| Li_2019 | 91 for sensitivity analysis, 19 for specificity analysis | 1 | 0 | 2 | 107 |
| Park_2021 | 262 | 3 | 0 | 12 | 247 |
| Sabari_2019 | 106 | 1 | 2 | 1 | 102 |

TP: true positive; FP: false positive; FN: false negative; TN: true negative

Figure S14. Forest plot of sensitivity and specificity on ctDNA detection of *ROS1* from bivariate random effects meta-analyses (n=4)

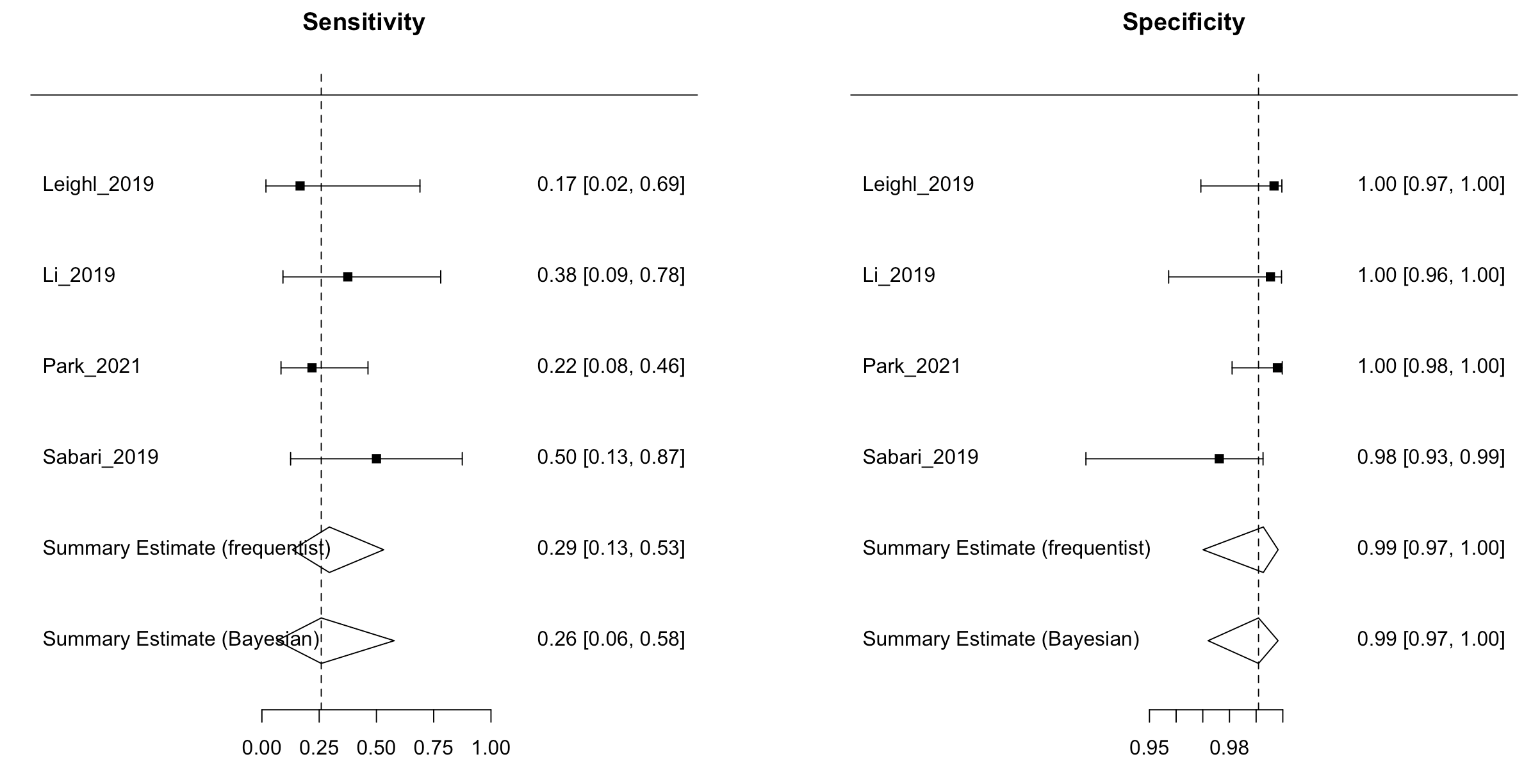

Sensitivity and specificity of single studies were based on frequentist estimations; I^2^ = 0% based on the frequentist approach

Figure S15. Summary receiver operating characteristics (SROC) plots based on bivariate random-effects meta-analyses of ctDNA detection of *ROS1* (n=4)

(b) SROC (frequentist approach)

(c) SROC (Bayesian approach):

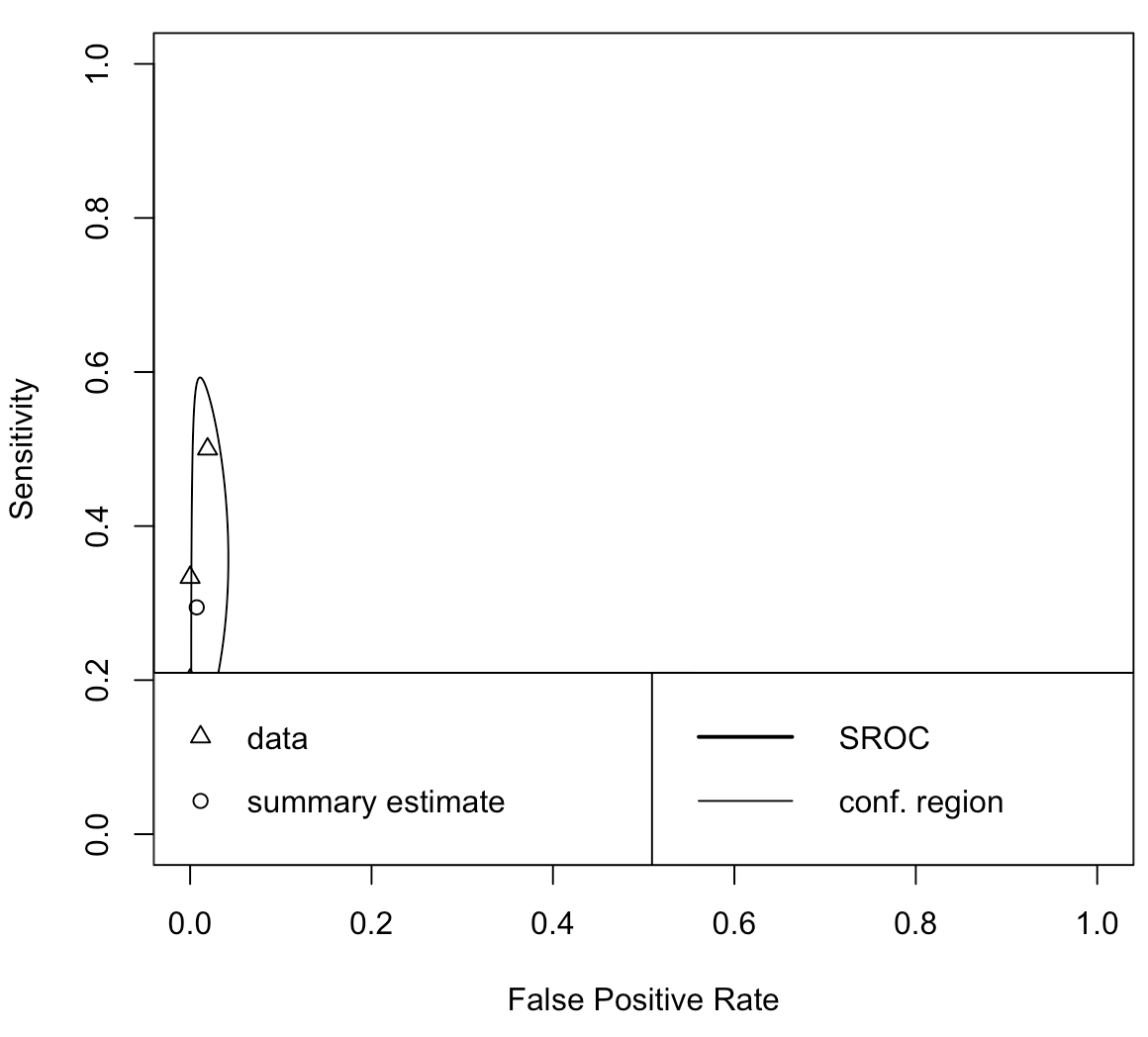

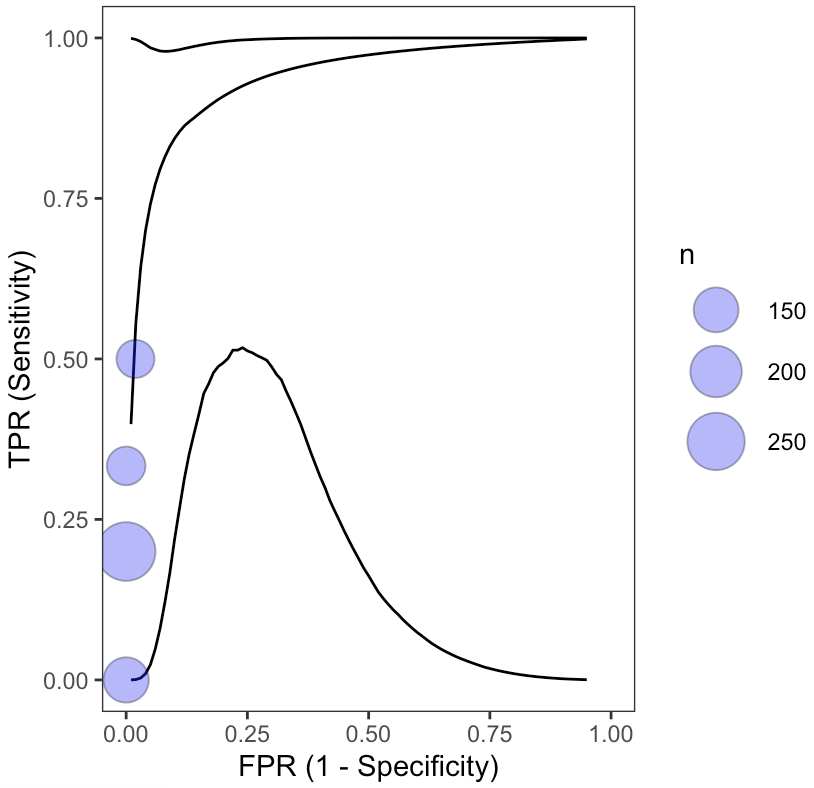

Figure S16. Bayesian Posterior Predictive Contours (50%, 75%, 95%) for ctDNA detection of *ROS1* (n=4)

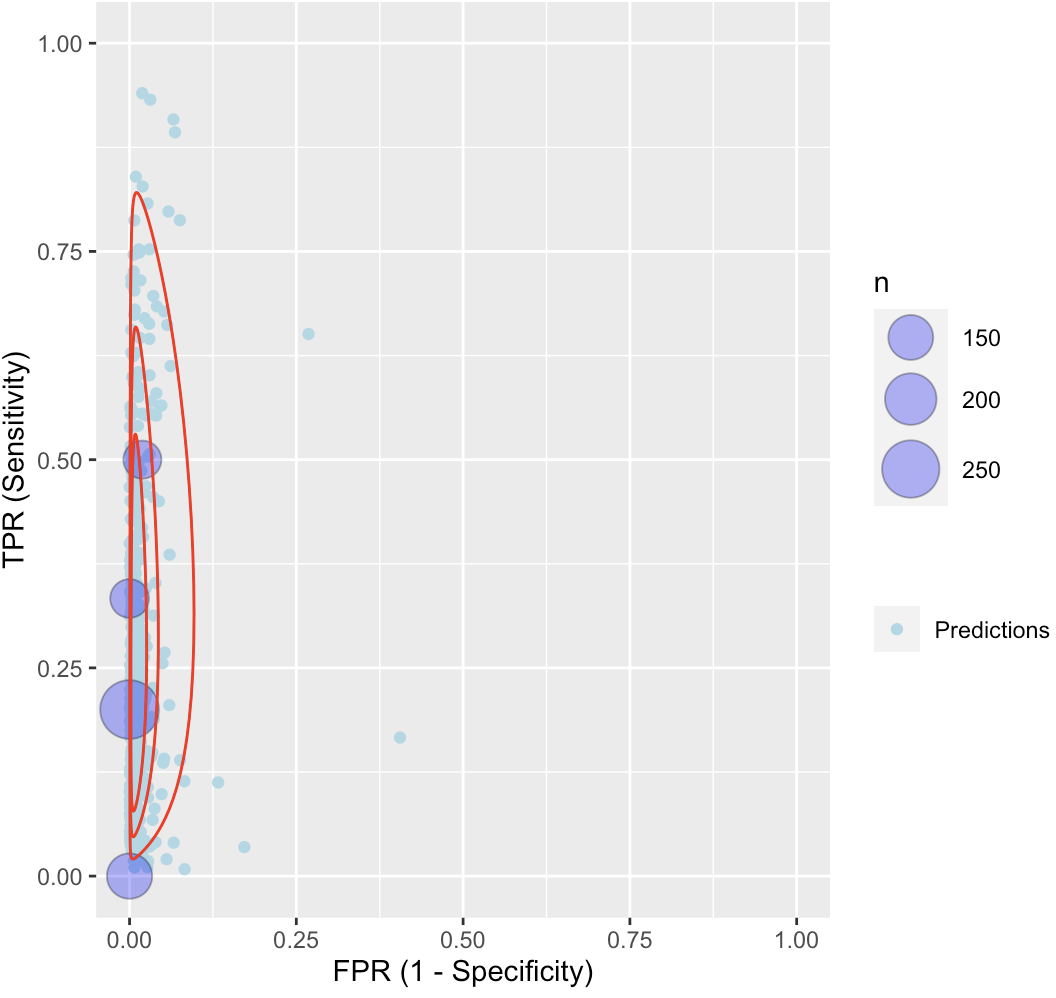

Each purple circle identifies the true positive rate (TPR) vs. the false positive rate (FPR) of each study (observed data). Light blue circles are predictions based on Bayesian meta-analysis of data by fitting a bivariate random effects model, with red lines indicating parametric predictive contours.

Table S11. Data included in meta-analysis on the CV of ctDNA detection of *MET* (n=5)

| **Author_Year** | **No. of patients** | **TP** | **FP** | **FN** | **TN** |
| --- | --- | --- | --- | --- | --- |
| Li_2019 | 91 for sensitivity analysis, 19 for specificity analysis | 3 | 0 | 3 | 104 |
| Park_2021 | 262 | 8 | 9 | 11 | 496 |
| Pritchett_2019 | 140 | 3 | 0 | 3 | 133 |
| Remon_2019 | 94 | 1 | 0 | 2 | 45 |
| Sabari_2019 | 106 | 3 | 0 | 1 | 102 |

TP: true positive; FP: false positive; FN: false negative; TN: true negative

Figure S17. Forest plot of sensitivity and specificity on ctDNA detection of *MET* from bivariate random effects meta-analyses (n=5)

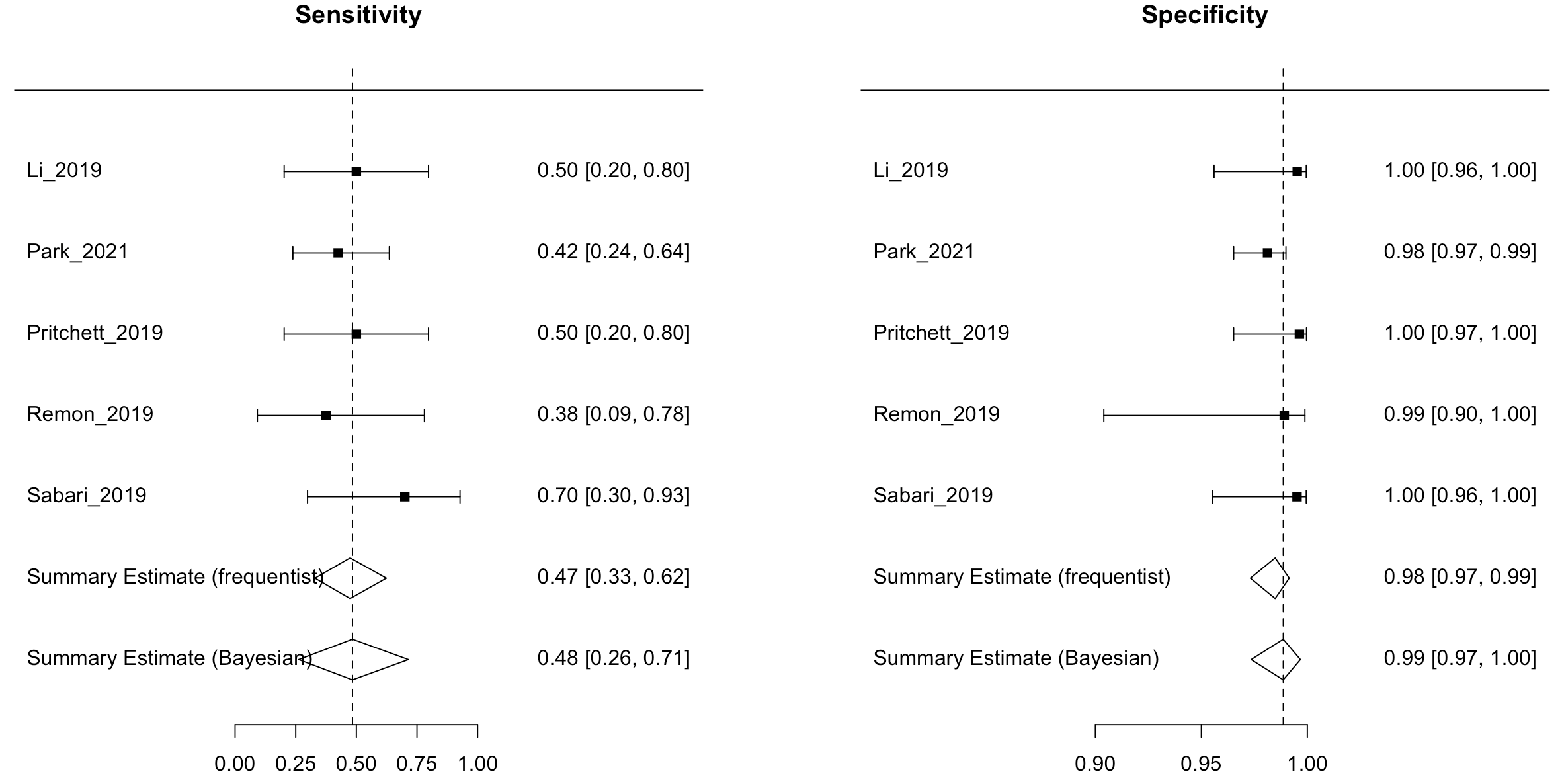

Sensitivity and specificity of single studies were based on frequentist estimations; I^2^ = 7.3% based on the frequentist approach

Figure S18. Summary receiver operating characteristics (SROC) plots based on bivariate random-effects meta-analyses of ctDNA detection of *MET* (n=5)

(a) SROC (frequentist approach):

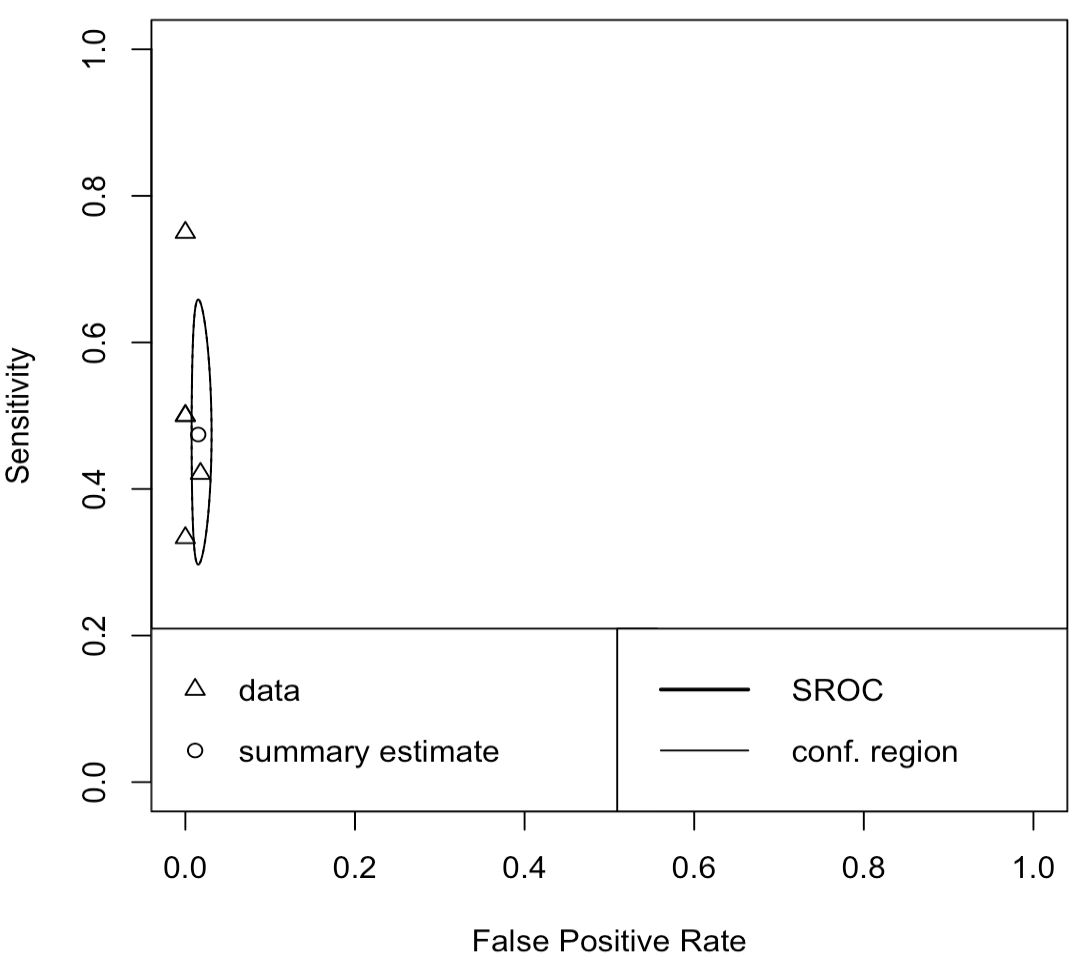

(b) SROC (Bayesian approach):

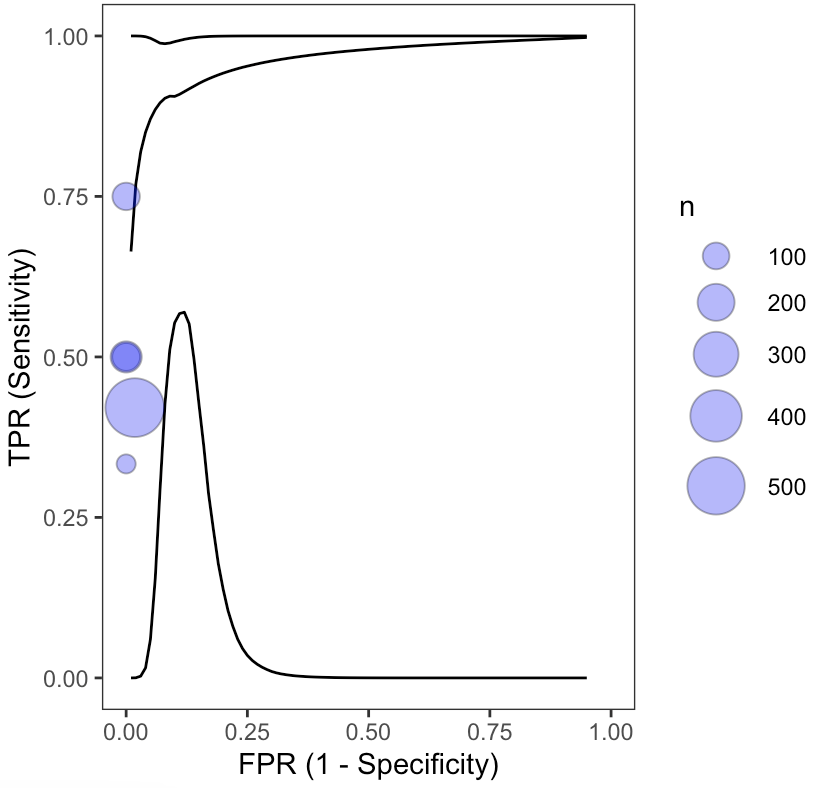

Figure S19. Bayesian Posterior Predictive Contours (50%, 75%, 95%) for ctDNA detection of *MET* (n=5)

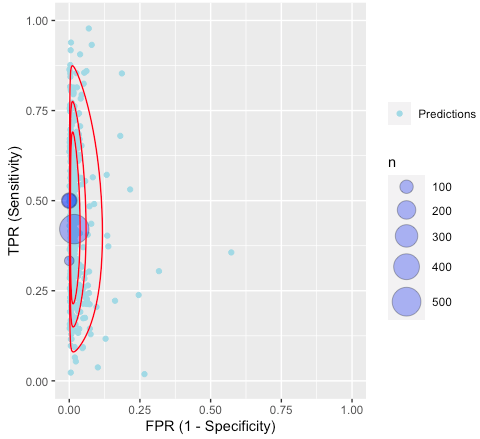

Each purple circle identifies the true positive rate (TPR) vs. the false positive rate (FPR) of each study (observed data). Light blue circles are predictions based on Bayesian meta-analysis of data by fitting a bivariate random effects model, with red lines indicating parametric predictive contours.

Table S12. Data included in meta-analysis on the CV of ctDNA detection of *RET* (n=3)

| **Author_Year** | **No. of patients** | **TP** | **FP** | **FN** | **TN** |
| --- | --- | --- | --- | --- | --- |
| Li_2019 | 91 for sensitivity analysis, 19 for specificity analysis | 0 | 0 | 1 | 109 |
| Park_2021 | 262 | 7 | 2 | 12 | 241 |
| Sabari_2019 | 106 | 1 | 1 | 1 | 103 |

TP: true positive; FP: false positive; FN: false negative; TN: true negative

Figure S20. Forest plot of sensitivity and specificity on ctDNA detection of *RET* from bivariate random effects meta-analyses (n=3)

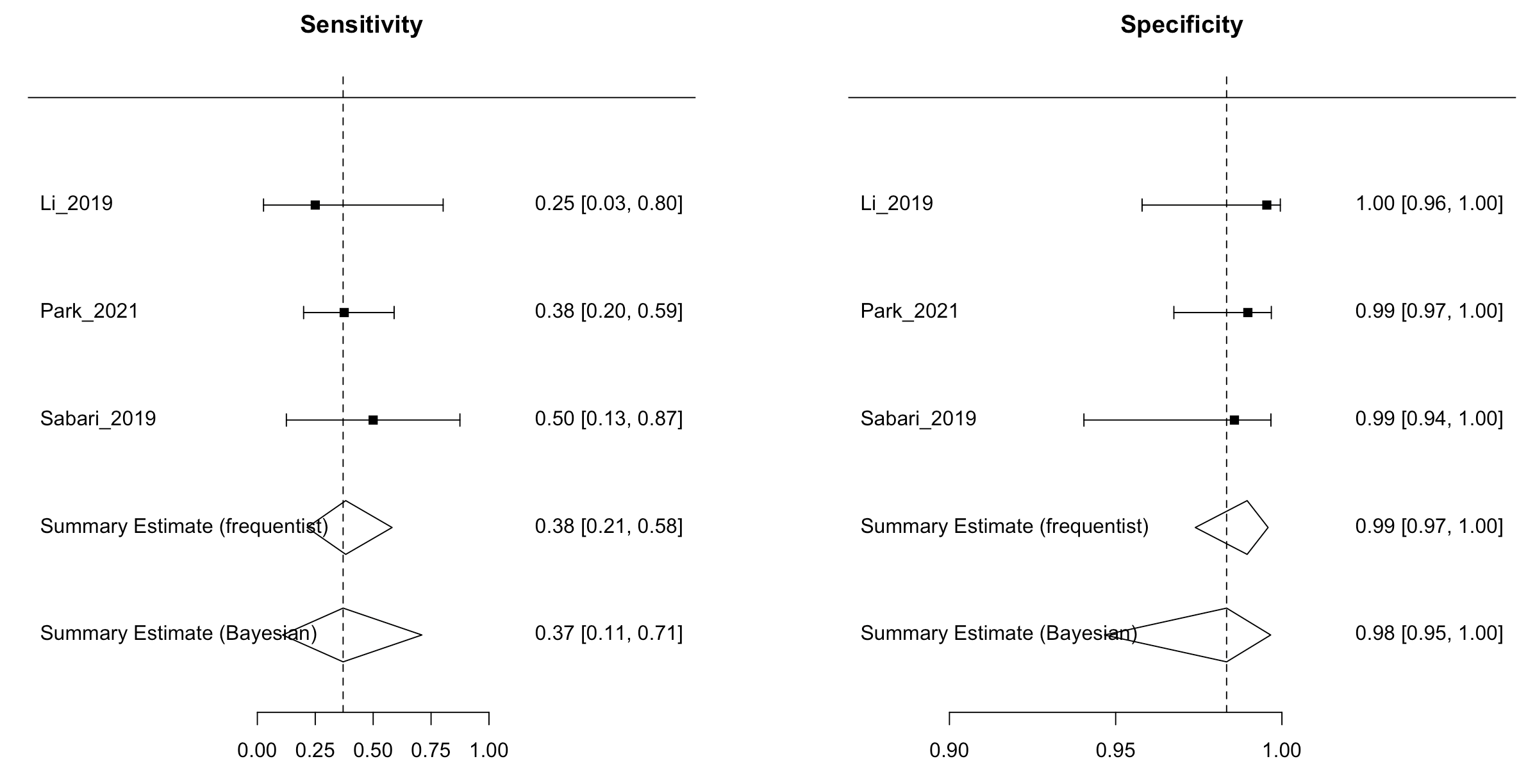

Sensitivity and specificity of single studies were based on frequentist estimations; I^2^ = 0% based on the frequentist approach

Figure S21. Summary receiver operating characteristics (SROC) plots based on bivariate random-effects meta-analyses of ctDNA detection of *RET* (n=3)

(a) SROC (frequentist approach):

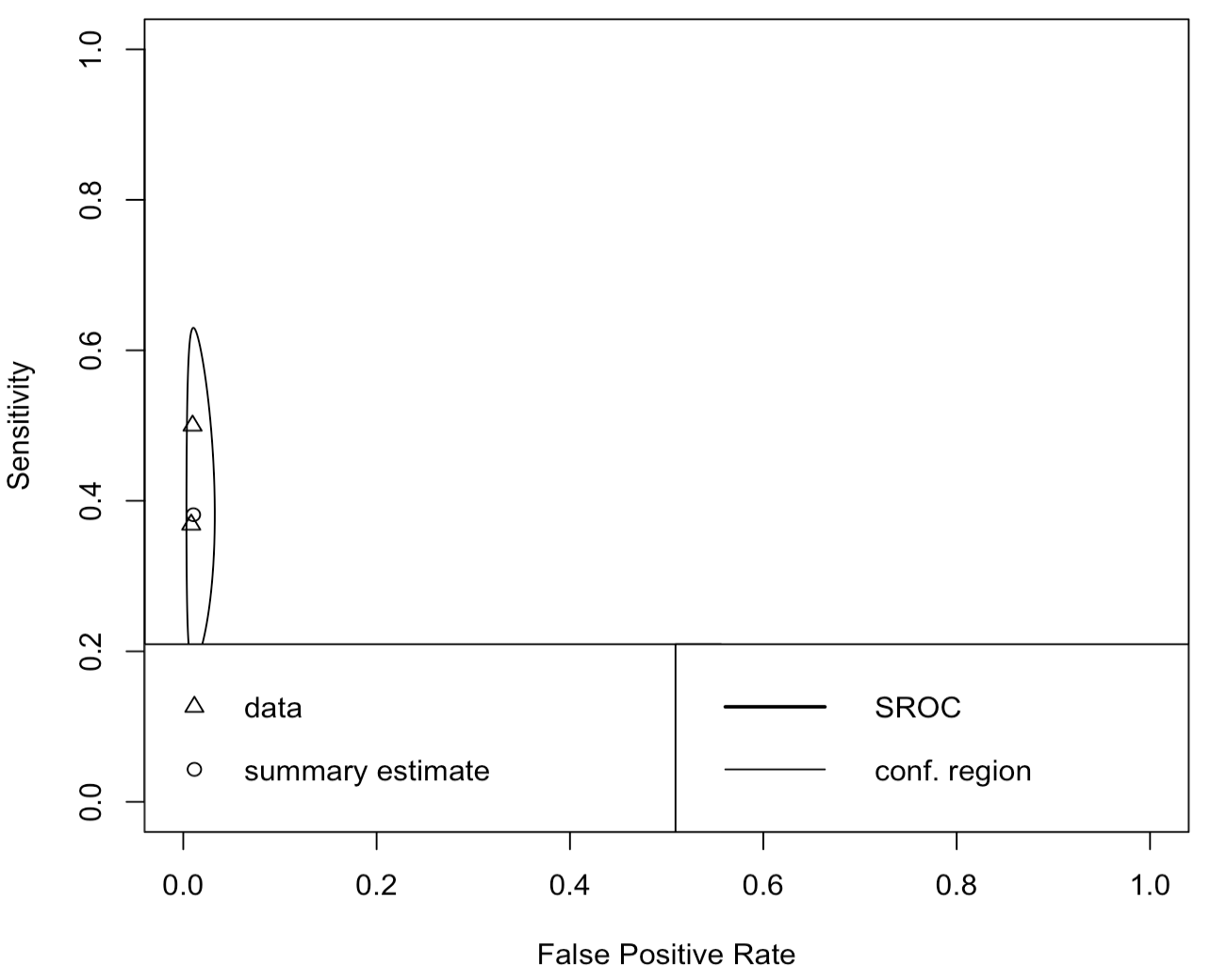

(b) SROC (Bayesian approach):

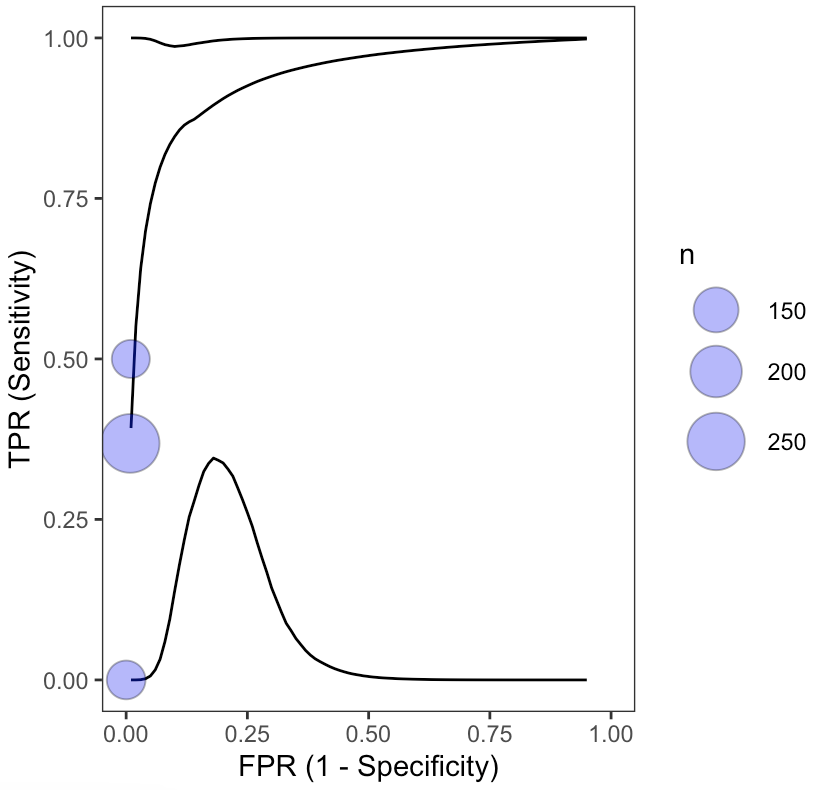

Figure S22. Bayesian posterior predictive contours (50%, 75%, 95%) for ctDNA detection of *RET* (n=3)

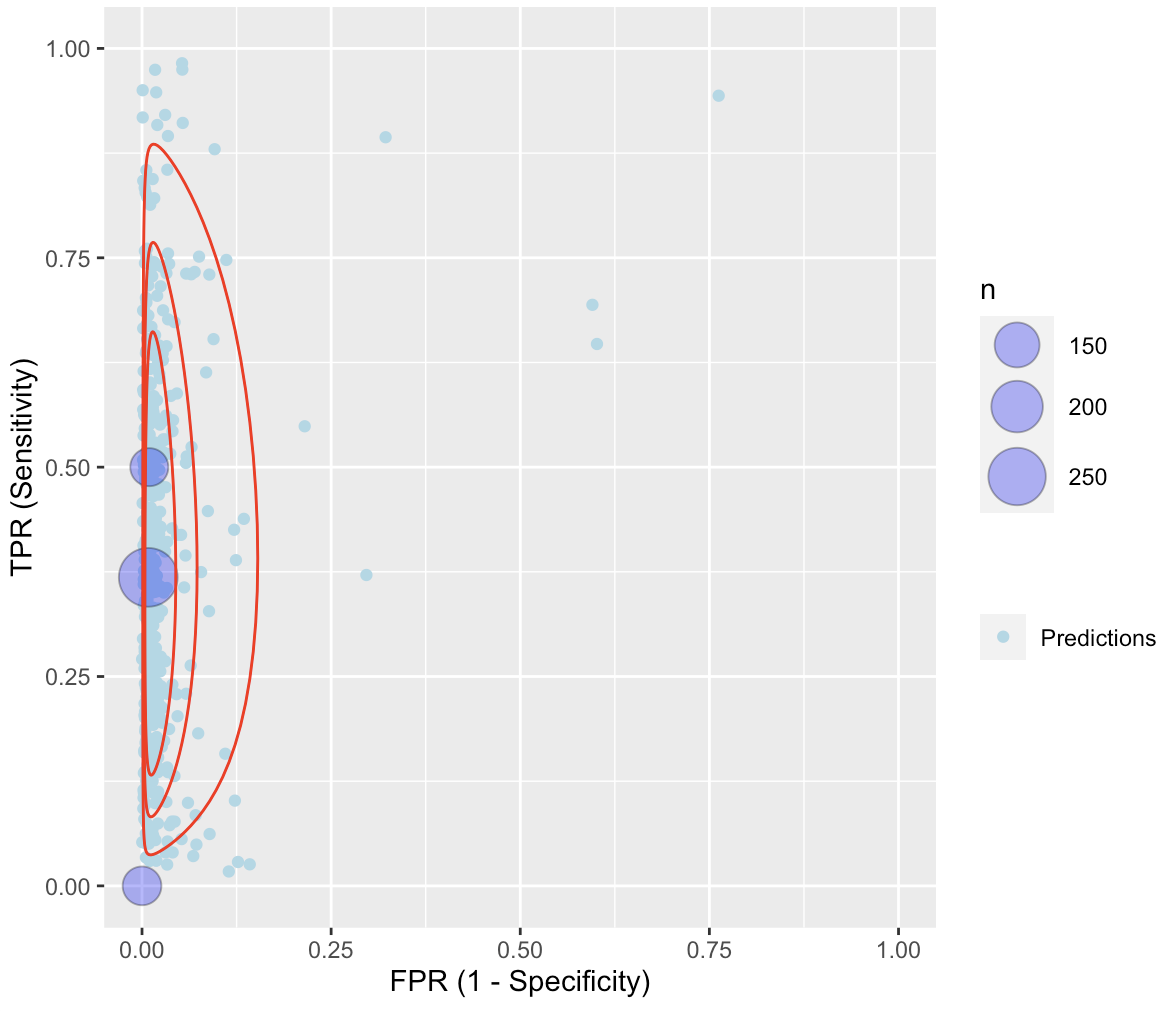

Each purple circle identifies the true positive rate (TPR) vs. the false positive rate (FPR) of each study (observed data). Light blue circles are predictions based on Bayesian meta-analysis of data by fitting a bivariate random effects model, with red lines indicating parametric predictive contours.

Table S13. Data included in meta-analysis on the CV of ctDNA detection of SNV mutation class (n=11)

| **Author_Year** | **No. of patients** | **TP** | **FP** | **FN** | **TN** |
| --- | --- | --- | --- | --- | --- |
| Leighl_2019 | 223 | 11 | 0 | 1 | 303 |
| Li_2019 | 91 for sensitivity analysis, 19 for specificity analysis | 26 | 0 | 6 | 188 |
| Müller_2017 | 29 | 4 | 0 | 6 | 48 |
| Park_2021 | 262 | 44 | 5 | 21 | 716 |
| Pritchett_2019 | 164 | 55 | 1 | 14 | 226 |
| Yao_2017 | 39 | 12 | 0 | 4 | 179 |
| Xu_2015 | 42 | 9 | 19 | 2 | 96 |
| Liu_2017 | 72 | 22 | 0 | 7 | 108 |
| Couraud_2014 | 45 | 2 | 0 | 3 | 168 |
| Chen_2019 | 7 | 1 | 0 | 0 | 4 |
| Remon_2019 | 94 | 23 | 7 | 4 | 131 |

TP: true positive; FP: false positive; FN: false negative; TN: true negative; SNV: single nucleotide variant or polymorphism, including 1) *EGFR* L858R, T790M, L861Q, G719S, and other unspecified *EGFR* SNVs, 2) *KRAS* G12X and other unspecified *KRAS* SNVs, 3) *BRAF* V600E and other unspecified *BRAF* SNVs

Figure S23. Forest plot of sensitivity and specificity on ctDNA detection of SNVs from bivariate random effects meta-analyses (n=11)

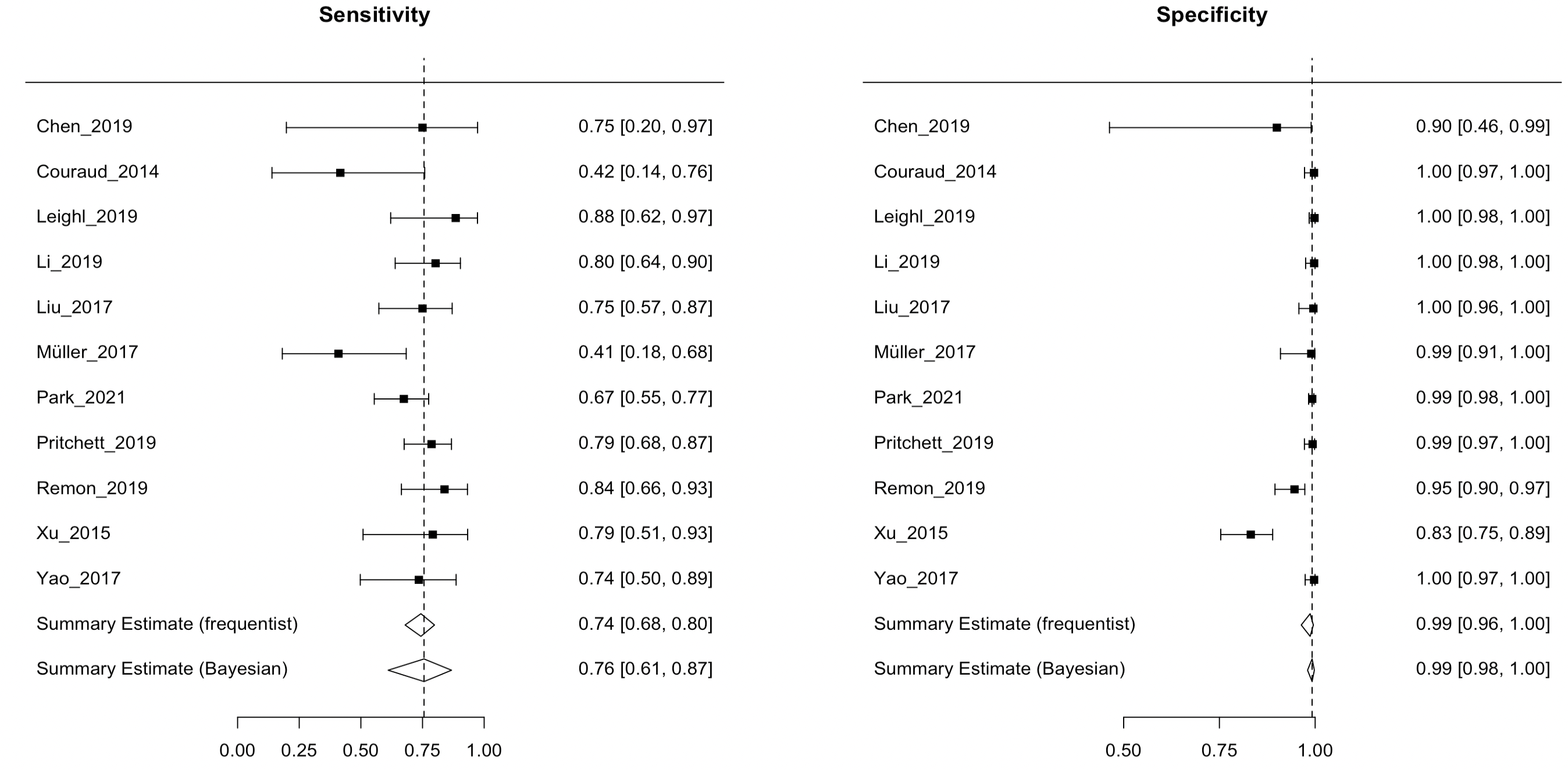

Sensitivity and specificity of single studies were based on frequentist estimations.

I^2^ = 0% based on the frequentist approach; BAUC = 0.72 (95%CI: 0.70-0.74)

Figure S24. Summary receiver operating characteristics (SROC) plots based on bivariate random-effects meta-analyses of ctDNA detection of SNV mutation class (n=11)

(a) SROC (frequentist approach):

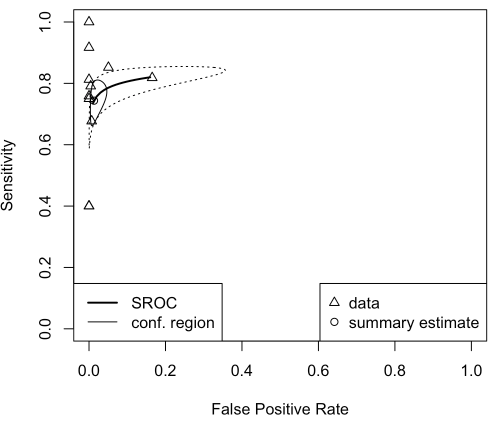

(b) SROC (Bayesian approach):

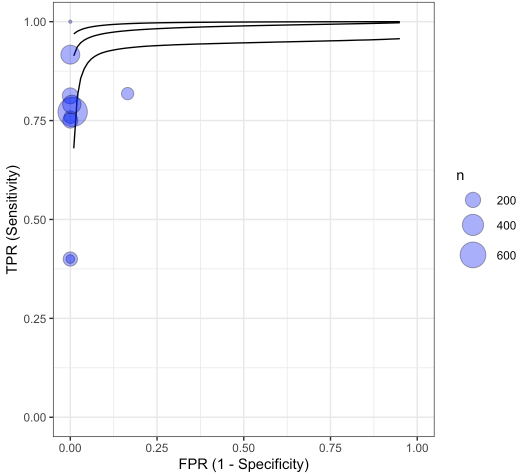

Figure S25. Bayesian posterior predictive contours (50%, 75%, 95%) for ctDNA detection of SNV mutation class (n=11)

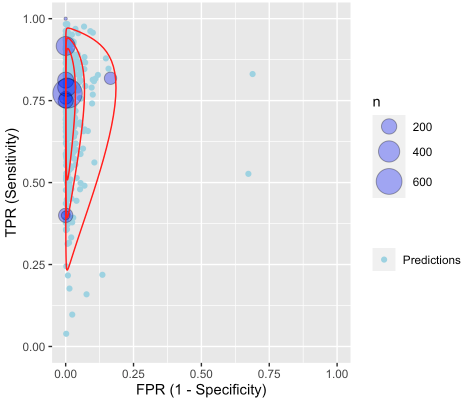

Each purple circle identifies the true positive rate (TPR) vs. the false positive rate (FPR) of each study (observed data). Light blue circles are predictions based on Bayesian meta-analysis of data by fitting a bivariate random effects model, with red lines indicating parametric predictive contours.

Table S14. Data included in meta-analysis on the CV of ctDNA detection of indel mutation class (n=6)

| **Author_Year** | **No. of patients** | **TP** | **FP** | **FN** | **TN** |
| --- | --- | --- | --- | --- | --- |
| Leighl_2019 | 223 | 18 | 0 | 4 | 201 |
| Park_2021 | 262 | 15 | 3 | 6 | 238 |
| Yao_2017 | 39 | 4 | 0 | 2 | 33 |
| Xu_2015 | 42 | 1 | 1 | 1 | 39 |
| Liu_2017 | 72 | 10 | 0 | 7 | 55 |
| Chen_2019 | 7 | 1 | 0 | 1 | 4 |

TP: true positive; FP: false positive; FN: false negative; TN: true negative; indels include *EGFR* exon 19 del or other unspecified *EGFR* indels

Figure S26. Forest plot of sensitivity and specificity on ctDNA detection of indels from bivariate random effects meta-analyses (n=6)

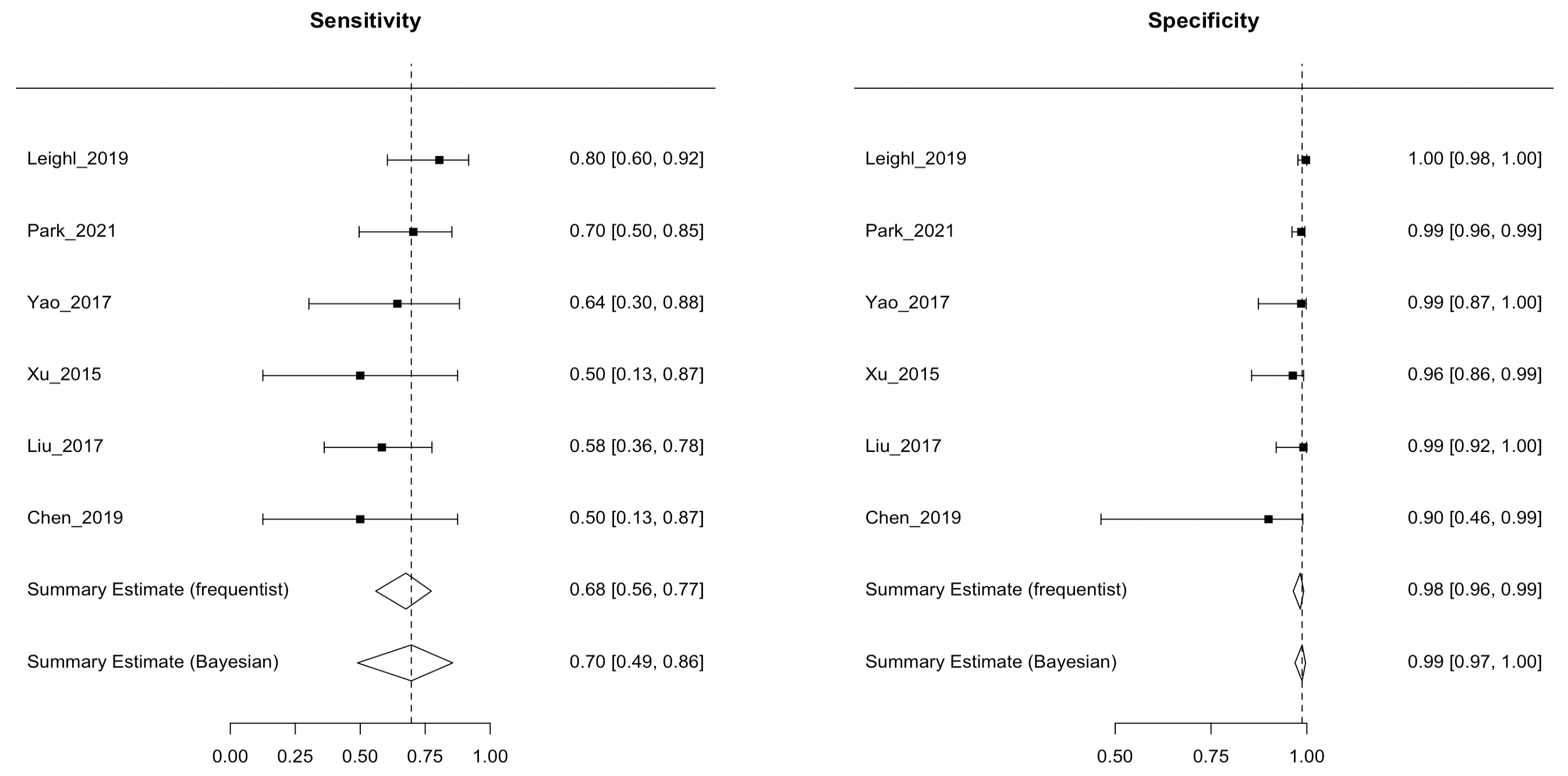

Sensitivity and specificity of single studies were based on frequentist estimations.

I^2^ = 3.3% based on the frequentist approach; BAUC = 0.52 (95%CI: 0.48-0.54)

Figure S27. Summary receiver operating characteristics (SROC) plots based on bivariate random-effects meta-analyses of ctDNA detection of indel mutation class (n=6)

(a) SROC (frequentist approach):

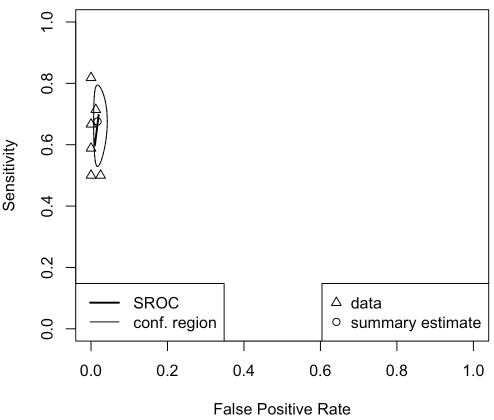

(b) SROC (Bayesian approach):

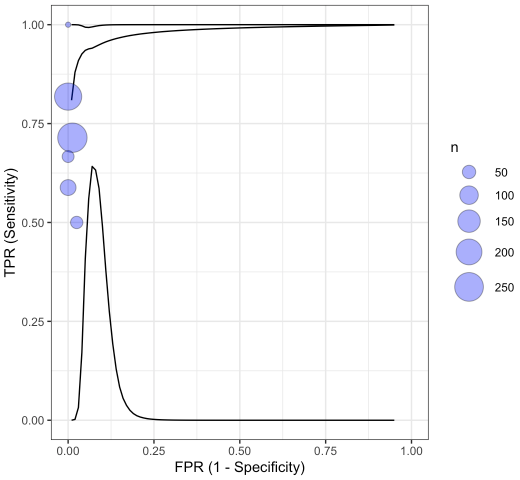

Figure S28. Bayesian posterior predictive contours (50%, 75%, 95%) for ctDNA detection of indel mutation class (n=6)

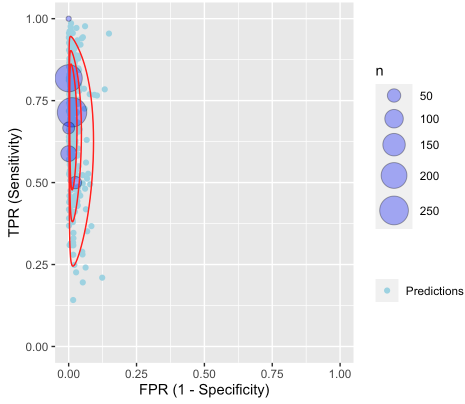

Each purple circle identifies the true positive rate (TPR) vs. the false positive rate (FPR) of each study (observed data). Light blue circles are predictions based on Bayesian meta-analysis of data by fitting a bivariate random effects model, with red lines indicating parametric predictive contours.

Table S15. Data included in meta-analysis on the CV of ctDNA detection of fusion mutation class (n=5)

| **Author_Year** | **No. of patients** | **TP** | **FP** | **FN** | **TN** |
| --- | --- | --- | --- | --- | --- |
| Leighl_2019 | 223 | 6 | 0 | 4 | 358 |
| Li_2019 | 91 for sensitivity analysis, 19 for specificity analysis | 6 | 0 | 6 | 318 |
| Liu_2017 | 72 | 2 | 1 | 2 | 59 |
| Palmero_2021 | 132 | 2 | 1 | 3 | 126 |
| Park_2021 | 262 | 16 | 2 | 28 | 740 |

TP: true positive; FP: false positive; FN: false negative; TN: true negative; fusions include ALK, ROS1 and RET

Figure S29. Forest plot of sensitivity and specificity on ctDNA detection of fusions from bivariate random effects meta-analyses (n=5)

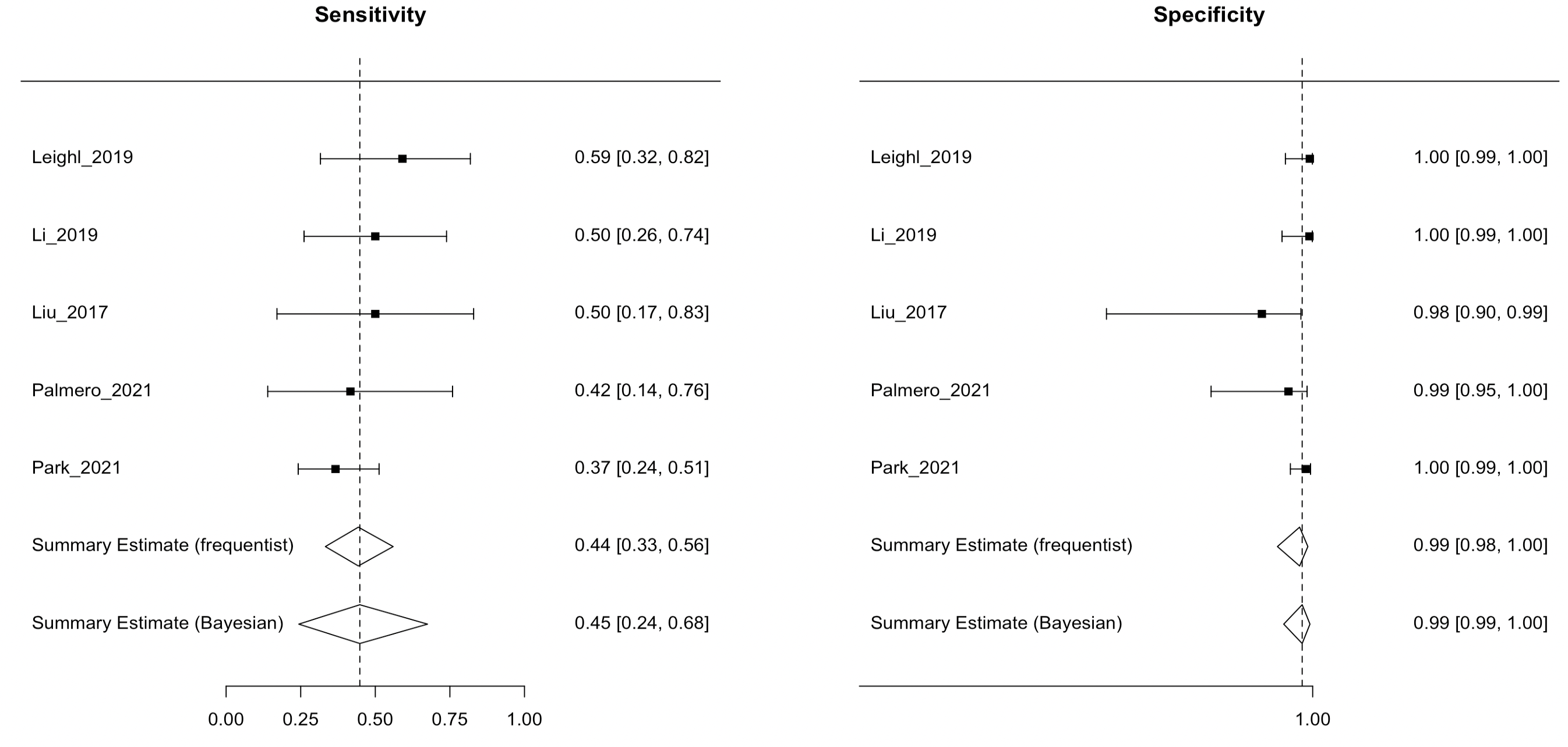

Sensitivity and specificity of single studies were based on frequentist estimations.

I^2^ = 0% based on the frequentist approach; BAUC = 0.52 (95%CI: 0.47-0.54)

Figure S30. Summary receiver operating characteristics (SROC) plots based on bivariate random-effects meta-analyses of ctDNA detection of fusion mutation class (n=5)

(a) SROC (frequentist approach):

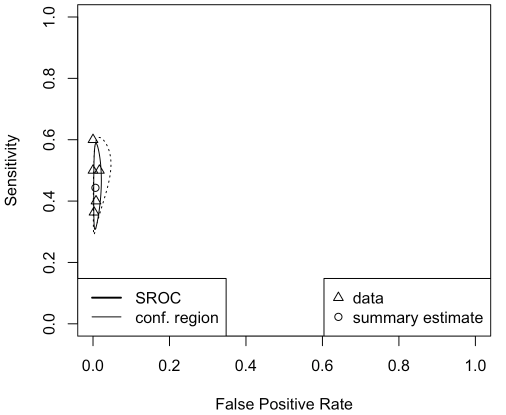

(b) SROC (Bayesian approach):

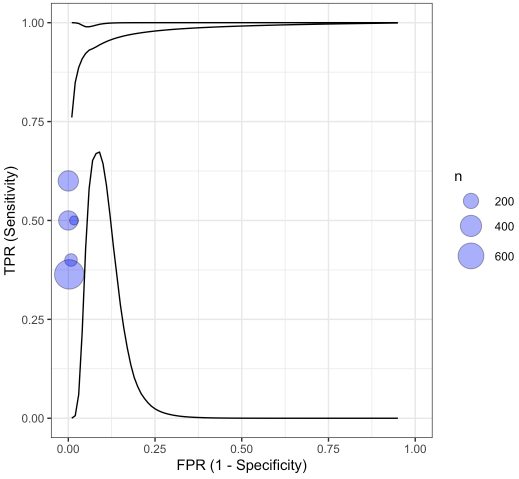

Figure S31. Bayesian posterior predictive contours (50%, 75%, 95%) for ctDNA detection of fusion mutation class (n=5)

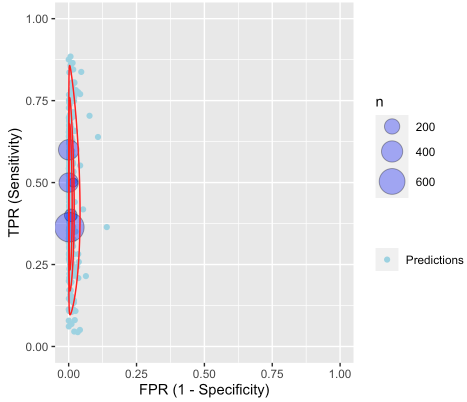

Each purple circle identifies the true positive rate (TPR) vs. the false positive rate (FPR) of each study (observed data). Light blue circles are predictions based on Bayesian meta-analysis of data by fitting a bivariate random effects model, with red lines indicating parametric predictive contours.

Table S16. Quality assessment of studies by QUADAS-2

| **Study ID** | **Risk of Bias** | | | | **Applicability Concerns** | | |
| --- | --- | --- | --- | --- | --- | --- | --- |
|  | Patient selection | Index Test(s) | Reference Standard | Flow and Timing | Patient selection | Index Test(s) | Reference Standard |
| Bustamante Alvarez_2021 | ☺ | ☺ | ☺ | ☹ | ☺ | ☺ | ☹ |
| Fernandes_2021 | ☺ | ☺ | ☺ | ☺ | ☺ | ☺ | ☺ |
| Leighl_2019 | ☺ | ☺ | ☹ | ☺ | ☺ | ☺ | ☺ |
| Li_2019 | ☹ | ☹ | ☺ | ☹ | ☺ | ☹ | ☺ |
| Lin_2021 | ☺ | ☺ | ☺ | ☹ | ☺ | ☺ | ☹ |
| Müller_2017 | ☺ | ? | ☺ | ☹ | ☹ | ☺ | ☺ |
| Palmero_2021 | ☺ | ☺ | ☺ | ☺ | ☺ | ☺ | ☺ |
| Park_2021 | ☺ | ☺ | ☺ | ☺ | ☺ | ☺ | ☺ |
| Pritchett_2019 | ☺ | ☺ | ☺ | ☹ | ☺ | ? | ☺ |
| Schwaederlé_2017 | ? | ☹ | ☹ | ☹ | ☹ | ? | ☺ |
| Yao_2017 | ☺ | ☺ | ☺ | ☺ | ☹ | ☺ | ☺ |
| Xu_2016 | ? | ? | ☺ | ☺ | ? | ? | ☺ |
| Liu_2018 | ☺ | ☺ | ☺ | ☺ | ☺ | ? | ☺ |
| Couraud_2014 | ☺ | ☺ | ☺ | ☺ | ☺ | ☺ | ☺ |
| Chen_2019 | ? | ? | ☺ | ☺ | ☹ | ? | ☺ |
| Remon_2019 | ☺ | ☺ | ☺ | ☺ | ☺ | ☺ | ☺ |
| Guo_2016 | ? | ? | ☺ | ☺ | ☹ | ? | ☺ |
| Sabari_2019 | ☺ | ☺ | ☺ | ☺ | ? | ☺ | ☺ |

☺Low Risk	☹High Risk	? Unclear Risk
